# First-in-human trial in Burkinabe adults and children of replication-competent, genetically attenuated PfSPZ-LARC2 Vaccine for malaria

**DOI:** 10.64898/2026.10.16.26365330

**Authors:** Sodiomon B. Sirima, Thomas L Richie, Alphonse Ouedraogo, Ben I. Soulama, Aminata R. Zou, Issa Nebié, KC Natasha, Nelly Camargo, Debashree Goswami, Jean Moïse T. Kabore, Nafissatou Kirakoya, Rolel K. Mbaiasra, Sem Ezinmegnon, Emilie S. Badoum, Sinndia Y. Ouedraogo, Noelie B. Henry, Jean B. Yaro, Thabo Mabuka, Tooba Murshedkar, Yonas Abebe, Diana Perez, Eric R. James, Nana Minkah, Ashley M. Vaughan, Matthew B. Laurens, B. Kim Lee Sim, Stefan H. I. Kappe, Alfred B. Tiono, Stephen L. Hoffman

**Author notes:** Corresponding authors. (S.L.H.); (S.H.I.K); (S.B.S.). These authors contributed equally to this work (Sodiomon B. Sirima, Thomas L Richie).

## Abstract

The World Health Organization has called for malaria vaccines with > 90% efficacy against *Plasmodium falciparum* (Pf) infection. The live, non-attenuated PfSPZ-CVac (chloroquine) vaccine provides the strongest protection against controlled human malaria infection (CHMI). Its 100% protective efficacy at 12 weeks against genetically distant Pf stems from a highly amplified immune stimulus. After inoculation, each invading sporozoite (SPZ) replicates up to 50,000 times within a hepatocyte. Over six days of development, these parasites express thousands of distinct proteins, presenting a vast and diverse array of antigens to the host immune system. The late liver stage parasites then rupture into the bloodstream, where they are killed by the co-administered drug, chloroquine. To engineer a Pf parasite that develops to the late liver stage like PfSPZ-CVac (chloroquine), but then arrests before transitioning to the blood stage, two genes required for transition from non-pathogenic liver to pathogenic blood stages were identified and deleted, creating late-liver stage arresting, replication-competent (LARC) PfSPZ. In a first-in-human, randomized, double-blind, placebo-controlled clinical trial, we assessed PfSPZ-LARC2 Vaccine in 75 malaria-exposed Burkinabe 1-to 48-year-olds. No vaccine-strain breakthrough blood-stage infections were detected; safety and tolerability profiles were similar in vaccinees and placebo recipients; and the highest immunogenicity was observed in the youngest recipients. Based on these findings, PfSPZ-LARC2 Vaccine has progressed to Phase 2 testing with the goal of rapidly advancing this genetically attenuated whole parasite vaccine to licensure and deployment.

**One Sentence Summary:** PfSPZ-LARC2 Vaccine, a replication-competent genetically attenuated malaria sporozoite vaccine, was safe and immunogenic in 1-48-year-old Burkinabes

## INTRODUCTION

In 2024, malaria caused 282 million clinical cases and 610,000 deaths, 6% more deaths and 23% more cases than in 2015, despite an annual investment in malaria control measures of ∼$4.5 billion (*1*). In 2023, there were more cases of malaria imported into the United States (US) than in any year since 1972, and in 2023 and 2025 malaria was transmitted in 6 states in the US, the first autochthonous transmission in two decades (*2*). In Europe, imported malaria cases have similarly increased (*3*). In endemic countries, the effectiveness of current control measures plateaued in about 2014. Since then, malaria’s impact in many countries, especially in sub-Saharan Africa, has remained the same or worsened. New, more effective measures are needed, such as high-efficacy vaccines.

*Plasmodium falciparum* (Pf), which causes more than 98% of all deaths from malaria, is a protozoan parasite that expresses more than 5,000 proteins (*4*) during its complex life cycle that could serve as vaccine immunogens. In the past few years, two moderately effective, subunit, recombinant protein malaria vaccines based on one parasite protein, Pf circumsporozoite protein (CSP), RTS,S-AS01 and R21-MatrixM, have been recommended for use by the World Health Organization in older infants and young children in Africa (*5*) and have achieved significant reductions in morbidity and mortality when administered to 5-17 month olds using a four dose regimen. However, the World Health Organization (WHO) has called for a vaccine that prevents infection in more than 90% of recipients of all ages, to enable more effective malaria control and eventual elimination (*6*).

The only vaccines that have achieved >90% vaccine efficacy (VE) against controlled human malaria infection (CHMI) or any VE against field-transmitted Pf infection in adults have been vaccines comprised of aseptic, purified, cryopreserved Pf sporozoites (SPZ) called PfSPZ vaccines (*7–15*), which expose the immune system to 1000s of parasite antigens. Immunization pre-conception of Malian women with three doses of 9×10^5^ PfSPZ of the early liver stage arresting, non-replicating, radiation attenuated PfSPZ Vaccine had 57% VE against Pf infection during pregnancy for 18-21 months after the last dose of vaccine with no intervening booster doses (*15*). Infants born to the mothers who were immunized pre-conception had 95.6% protection against clinical malaria during their first year of life (*16*). The same immunization regimen gave 79% VE in non-immune adults against heterologous CHMI (challenge strain genetically distant from the vaccine strain) at 9-10 weeks after the last dose (*12*).

The strength and breadth of the immune stimulus provided by PfSPZ Vaccine can be increased by immunizing with non-attenuated parasites that replicate up to 50,000-fold and express ∼3,000 new proteins in the liver before being killed by a co-administered antimalarial drug (*8, 17*). This approach, called chemoprophylaxis vaccination (PfSPZ-CVac) when using purified, cryopreserved, non-attenuated PfSPZ (PfSPZ Challenge) and chloroquine as the co-administered drug, provided 100% and 77% VE against CHMI with heterologous Pf 12 weeks after immunization using 22% (*11*) and 12% (*18*) of the PfSPZ Vaccine dose that gave 79% VE at 9-10 weeks (*12*). Further development of the more potent PfSPZ-CVac is unlikely, however, due to the safety risks associated with relying on a co-administered drug for attenuation.

To create a replication-competent parasite like PfSPZ Challenge that amplifies antigen biomass and diversity but prevents the risk of breakthrough to blood-stage infection, we made PfSPZ-LARC1 (*19*), aka GA2 (*20*), by deleting the *Mei2* gene. This parasite had the desired phenotype – replication-competent, late liver-stage arresting (LARC), but when testing the *P. yoelii* (Py) rodent malaria ortholog, about 10% of mice immunized with as few as 4×10^4^ PySPZ developed blood-stage infections, indicating incomplete attenuation (*21*). In an effort to improve safety, we made PySPZ-LARC2 (2 = two deletions) by deleting *Linup* in addition to *Mei2*, and demonstrated that 5-fold higher inoculations (2×10^5^ PySPZ) of the doubly-deleted PySPZ were attenuated in all mice without any loss in protective efficacy (*21*). This led to the selection of PfSPZ-LARC2 for clinical development, reasoning that since each deletion independently resulted in a LARC phenotype, the combined genetic alterations would provide extra assurance of safety (*21*). The LARC2 parasite was manufactured in compliance with Good Manufacturing Practices as PfSPZ-LARC2 Vaccine. In mice with humanized livers, LARC2 parasites invade hepatocytes like wild-type PfSPZ, replicate, grow to a size equal to that of mature hepatic schizonts, but arrest development, disintegrating in the liver without transitioning to blood-stage infection, even after administering 1×10^6^ PfSPZ (*21*).

To test our hypothesis of complete attenuation and perform a preliminary assessment of safety, tolerability, and immunogenicity, we conducted a first-in-human, randomized, double-blind, placebo-controlled trial of PfSPZ-LARC2 Vaccine in a malaria-endemic region of Burkina Faso. A malaria-exposed population was selected because preexisting immunity, exposure risk, and safety considerations differ substantially from nonendemic settings. In addition, it was deemed appropriate to directly assess the vaccine in the intended use population. The study was initiated in adults, first demonstrating attenuation, safety, and tolerability in this non-vulnerable age group, then progressed to adolescents, school children, and pre-school children. This study, which began in April 2025, represents, to our knowledge, the first, first-in-human vaccine trial ever conducted in sub-Saharan Africa.

## RESULTS

### Study Groups

The study had 5 study groups divided into 3 cohorts. Cohort 1 was composed of adults (20-48 years) who received PfSPZ-LARC2 Vaccine in three doses of 2×10^5^ PfSPZ at four-week intervals (group 1) or one higher dose of 4×10^5^ PfSPZ (group 2). Cohort 2 was composed of adolescents (11-19 years) receiving one dose of 2×10^5^ PfSPZ (group 3), and school children (6-10 years) receiving one dose of 1×10^5^ PfSPZ (group 4). Cohort 3 was composed of pre-school children (1-5 years) receiving one dose of 1×10^5^ PfSPZ (group 5). The ten vaccinees and five normal saline placebo controls in each group were divided into a pilot group (2 vaccinees, 1 control) and a main group (8 vaccinees, 4 controls) and randomized 2:1 to PfSPZ-LARC2 Vaccine or normal saline placebo. Pilot groups initiated 3 days (groups 1-3) or 7 days (groups 4-5) before the main group to identify any concerning adverse events. In cohorts 1 and 2, the second pilot group was initiated three days after the first pilot group.

### Screening and Enrollment

Numbers screened and enrolled for groups 1-2 (screened together) and groups 3, 4 and 5 (each screened separately) were, 72/30, 26/15, 23/15, and 30/15, respectively (151/75 in total) (**Fig. 1**). All groups were fully enrolled. Demographic data are provided in **table S1**. Seventeen of the 76 participants screened but not enrolled were deemed eligible and kept as back-ups. Reasons for exclusion varied among groups (**Fig. 1**) but most common were positive serology for HIV, HBV or HCV, abnormal hematology or chemistry test results, electrocardiogram abnormalities (primarily adults), and body mass index (BMI) outside the 18-30 range. Thick blood smears done at the time of screening were positive in 0, 2 (13.3%), 8 (53.3%), and 0 enrolled participants in groups 1-2, 3, 4, and 5, respectively.

**Fig. 1.**
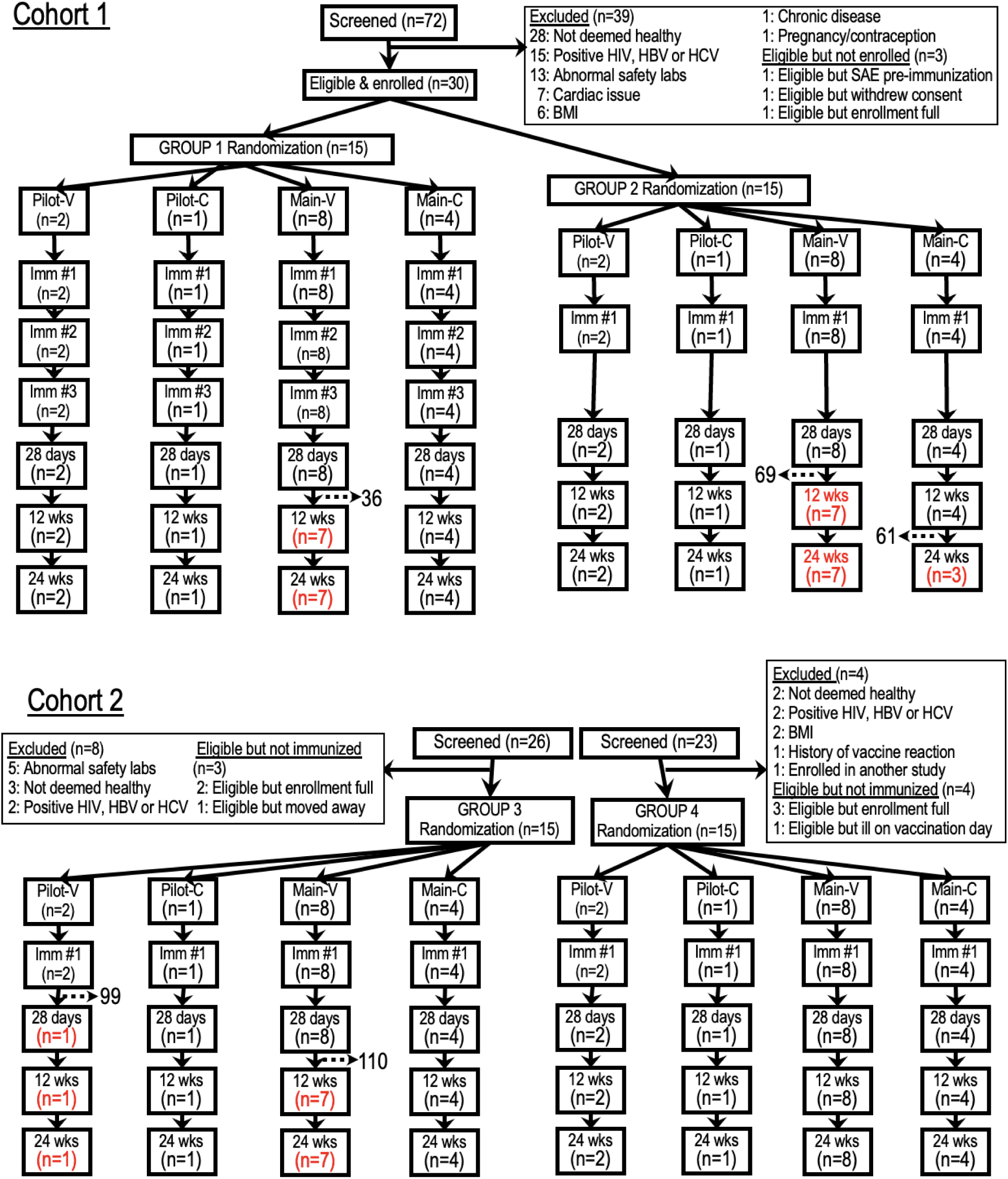
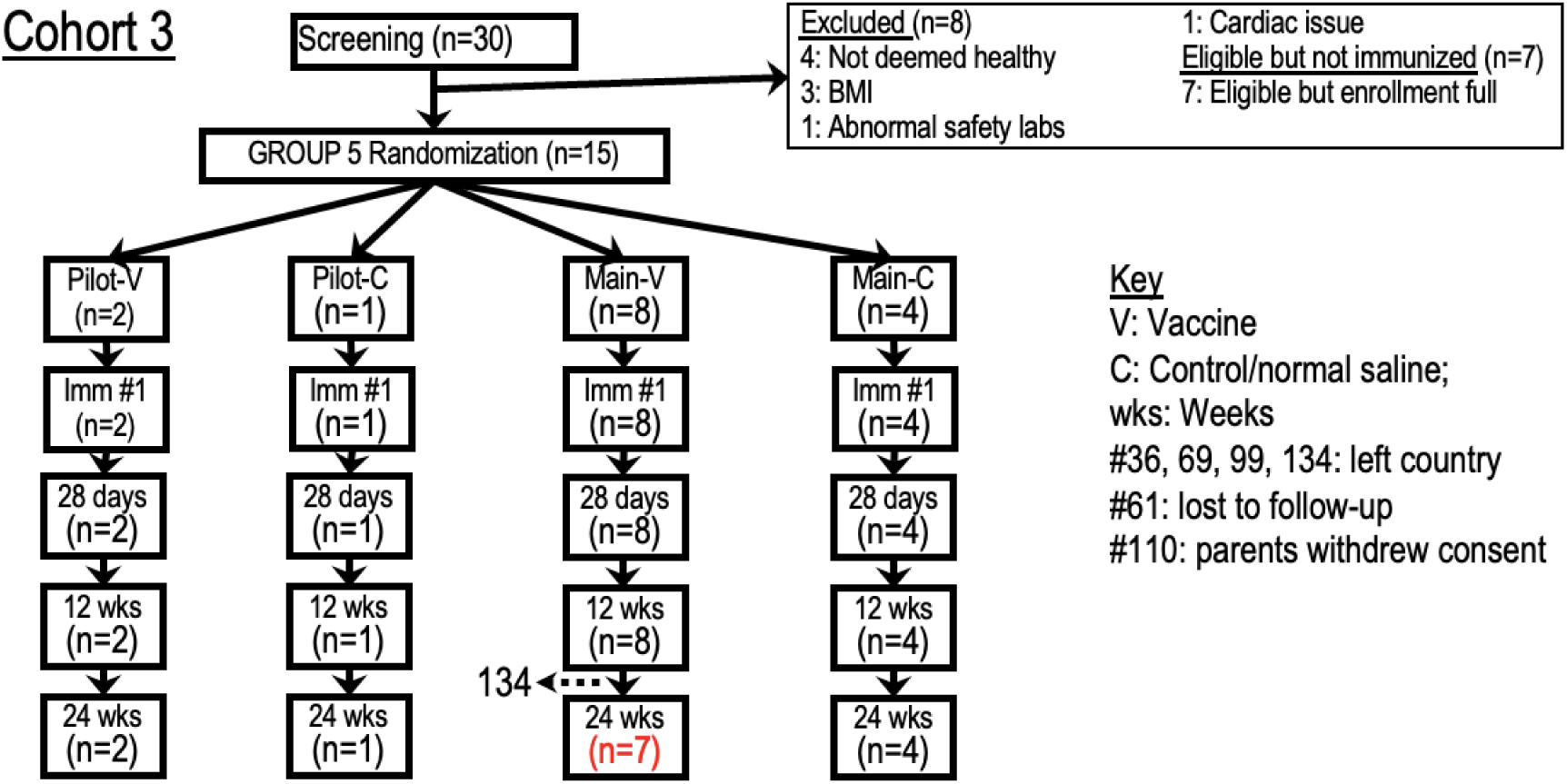
Participant flow. Participants were enrolled into three sequential cohorts by age and dose level. Flow diagram shows numbers of participants screened, excluded, enrolled, randomized, vaccinated, and completing follow-up. Dropouts are indicated by horizontal dashed lines and the participant ID number; subsequent diminished sample sizes are in red font. See text for discussion of dropouts. Several participants are counted under more than one reason for exclusion.

### Presumptive Pre-Treatment and Terminal Treatment

To clear any existing parasitemias that would otherwise confound the identification of vaccine strain breakthroughs, a three-day treatment course of artemether-lumefantrine was administered before immunization. This drug combination was selected because of the relatively shorter half-life of lumefantrine compared to other artemisinin combination therapies. Twelve days later, all participants were immunized (**Fig. 2**). Starting 28 days after immunization, all participants were terminally treated with dihydroartemisinin-piperaquine (DHA-P) for three days as a safety measure to eliminate any residual LARC2 parasites (**Fig. 2**).

**Fig. 2.**
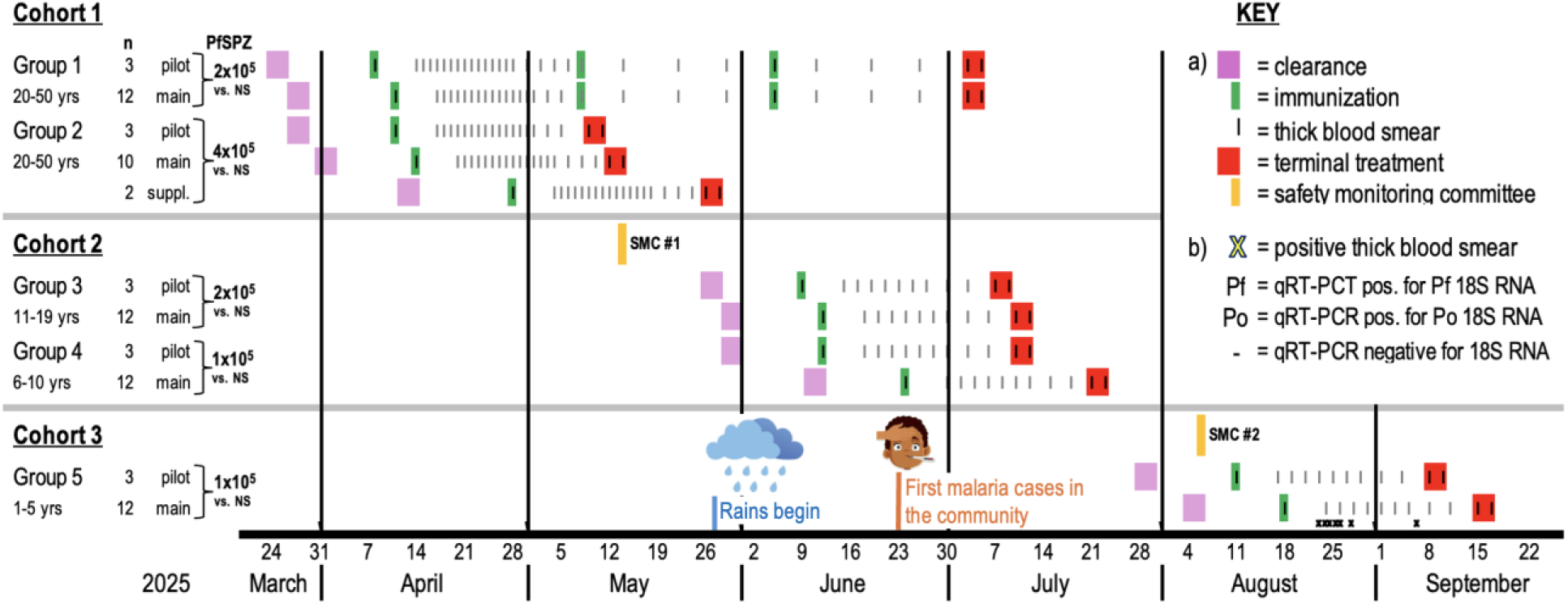
Trial execution. Three cohorts of healthy participants were cleared of any existing parasitemias (purple shading), immunized (green shading), followed clinically and by thick blood smear (tick marks), and received terminal treatment starting on day 28 after immunization (red shading). The Safety Monitoring Committee (SMC) reviewed existing data before recommending immunization of the next cohort (orange shading). There were no malaria infections diagnosed in cohorts 1 or 2 during 28 days of follow-up. There were seven infections diagnosed by thick blood smear in cohort 3 (small x’s above the timeline – see Fig. 3 for details). Group 2 supplement (suppl.): two additional participants were cleared to replace those who were withdrawn before immunization (see text).

### Study Flow

All participants completing initial pre-immunization drug clearance were immunized except for one consent withdrawal and one exclusion, both in group 2, the latter due to hospitalization for presumed appendicitis. Two replacements were cleared and immunized as a supplemental cohort (**Fig. 2**). During the remainder of the trial, 74/75 (98.7%) immunized participants completed 28-day follow-up and terminal treatment, and 69/75 (92%) completed the full study including 12- and 24-week follow-ups. One post-immunization drop-out was due to withdrawal of parental consent, and the other 5 were losses to follow-up, four because of travel to other regions or countries (see **supplementary material**).

### Assessment for vaccine breakthrough infections

Adult participants were assessed starting day 6 (5 days after immunization) by daily questioning about malaria symptoms, and, on days 6-20, 22, 24, 28 and 30, by measuring body temperature and reading a thick blood smear (TBS) (**Fig. 2**). When no parasitemias were documented in the 30 participants in cohort 1, the safety monitoring committee (SMC) reviewed the blinded safety data and recommended trial continuation. Cohort 2 (groups 3 and 4) was immunized and followed as above for signs and symptoms of malaria but with a reduced frequency of TBS to minimize needle sticks in this pediatric population: every other day from days 6-18 and days 21, 24, 28, and 30. When no parasitemias were detected in the 30 participants in cohort 2, the SMC reviewed the updated blinded safety data and recommended trial continuation. Cohort 3 (group 5) was immunized August 11-18 and followed with the same schedule as cohort 2, noting that the first rains were 28 May and malaria infections were diagnosed in the community by 23 June.

No infections were detected within 28 days of immunization in cohorts 1 and 2, which were immunized during the dry season and early rainy season. Seven infections were detected by thick blood smear in cohort 3 (group 5) (**Fig. 3A**). All confirmed infections were asymptomatic. Six were confirmed as malaria infections by a PCR assay (Pf, n=5; *P. ovale*, n=1), whereas one case was negative for *Plasmodium* by two different molecular assays (**Fig. 3B, C**), indicating a false-positive thick blood smear.

**Fig. 3.**
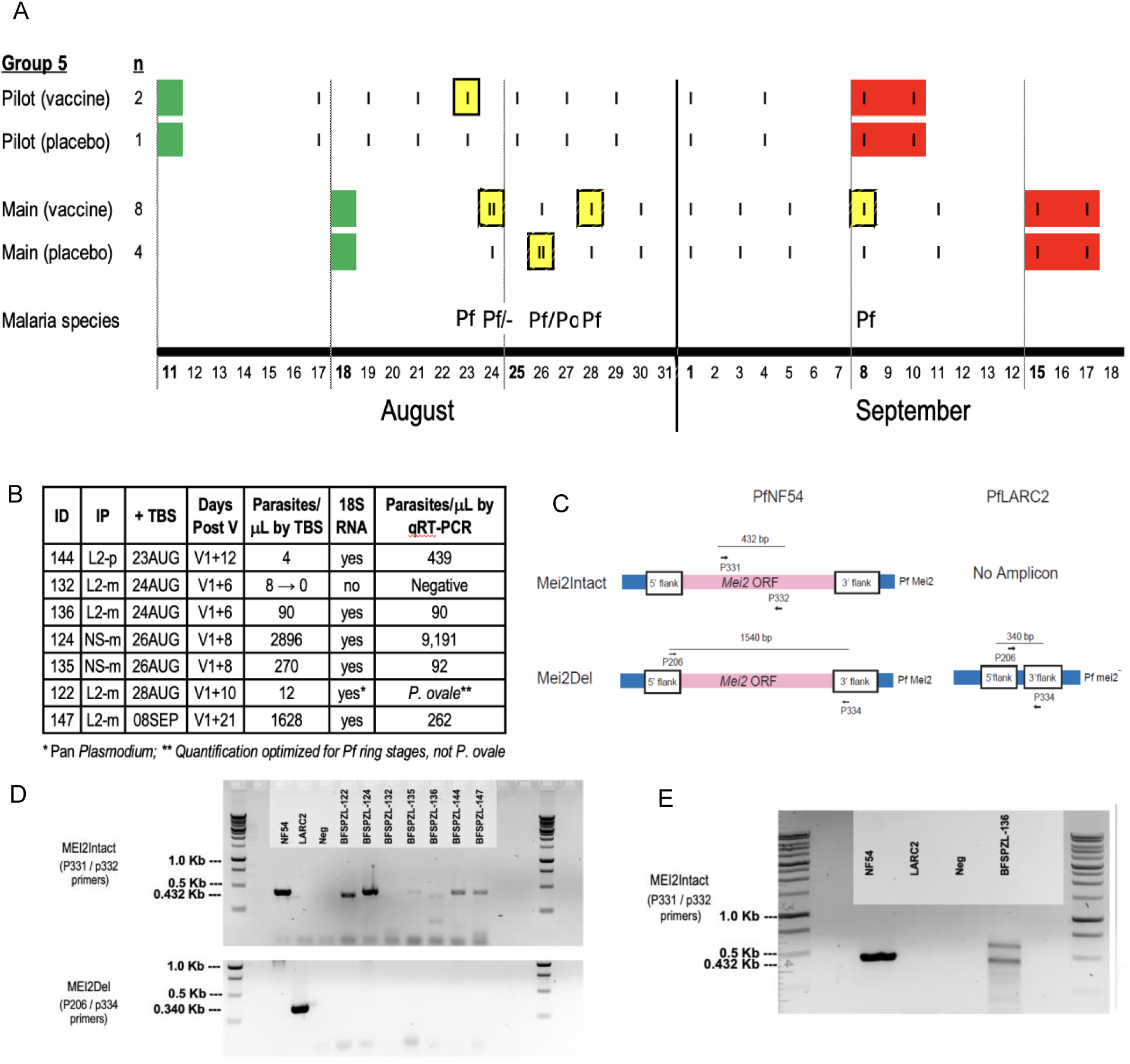
Assessment for breakthrough blood-stage infections in group 5. **A.** Seven infections were identified by TBS in group 5 and treated the same day, on the dates indicated, including two on Aug 24 and two on Aug 26 (yellow squares). Species are identified next to x-axis. **B.** Dried blood spots (DBS) were collected pre-treatment, total nucleic acids (TNA) eluted from the filter paper at the University of Washington, and assayed by qRT-PCR amplification of 18S RNA as a biomarker for *Plasmodium* blood-stage infection (*22*). This detected *Plasmodium* RNA in six samples, while the sample from BFSPZL-132 was negative, a result later confirmed using a second DBS. **C.** A PCR-based assay developed by SCRI employed two primer pairs to ascertain the identity of the Pf isolates: (1) The “*MEI2*Intact” primers p331/p332 bound internally to the *MEI-2* gene and generated a 432 base pair amplicon from parasites with the intact gene (wild type) and no amplicon from LARC2 parasites (vaccine strain); (2) The “*MEI2*Del” PCR primers p206/p334 bound externally to invariant *MEI-2* flanking regions to generate a large (1540 bp) amplicon from *MEI-2* intact parasites (wild-type) and a small (340 bp) amplicon from LARC2 parasites, noting, however, that the *Mei2*Del PCR was not optimized to amplify the larger band of 1540 bp in PfNF54. **D.** TNA from the seven samples was used in the identity assay. The *MEI2*Intact reaction generated amplicons of 432 bp, confirming the presence of the *MEI-2* gene in the NF54 (wild-type) control and the specimens from participants 124, 135, 144 and 147, while participant 132 had no product. Other results were deemed inconclusive: participants 122 and 136 showed bands smaller than predicted and required further study. The band from 122 was sequenced and matched the *MEI-2* sequence of *P. ovale* (**Fig. S1**). The *MEI2*Del reaction was also performed: only the LARC2 control generated the 340 bp amplicon, indicating absence of *MEI-2*. **E.** A second DBS from 136 was used to obtain additional TNA, and the *MEI2*Intact reaction showed two bands, the smaller one 432 pb in size. Both were sequenced, and the lower band matched Pf*MEI-2* sequence (**Fig. S2**). In conclusion, 122 was positive for *P. ovale,* and 124, 135, 136, 144, and 147 were positive for *MEI-2*-intact Pf, representing wild-type Pf parasites acquired through natural mosquito transmission. 132 was negative for *Plasmodium*. To reconcile this last result, which was discordant with the thick blood smear, the original smear was placed in a blinded set with others and read independently by 12 microscopists. Four read it as positive (median of 4 parasites/mL), 8 as negative. According to the GRAS SOP, the smear was re-scored as negative based on the majority reading (altered result for 132 in the table in panel **B**).

A genotyping assay demonstrated the presence of intact *MEI2* in all confirmed infections, indicating naturally acquired, wild-type parasites rather than vaccine parasites (**Fig. 3C-E**). In summary, no vaccine strain breakthrough blood-stage infections were detected in any participant.

### Safety and tolerability

Safety and tolerability were assessed by reviewing at each clinic visit (1) prespecified solicited local symptoms and signs referable to the site of injection (local AEs), (2) prespecified solicited systemic symptoms and axillary temperature (systemic AEs), (3) unsolicited AEs, identified by open-ended questioning, and (4) laboratory testing days 7 and 28 after immunization. All AEs and laboratory abnormalities (LAs) were rated by severity, unsolicited AEs for relatedness to immunization, and LAs for clinical significance (**supplementary material**).

PfSPZ-LARC2 was well tolerated by these measures across all dose levels and age groups. Solicited local AEs were uncommon, predominantly mild (grade 1), and similar in frequency between vaccine and placebo recipients (**Table 1**). Solicited systemic AEs occurred in 9/50 (18%) vaccine recipients and 3/25 (12%) placebo recipients (p=0.74), with no consistent dose- or age-related pattern (**Table 1**). Moderate (grade 2) AEs were infrequent and limited to adults, and no severe (grade 3) solicited systemic AEs were observed.

**Table 1.** Adverse events and laboratory abnormalities. Number (percent) of participants experiencing an AE or LA are provided in the global summary at the top of the table and the first two data columns. Data columns to the right of the first two columns provide the participant number experiencing an AE or LA by group (and severity grade), with superscripts denoting after which vaccination the AE or LA occurred for group 1 (all other groups received a single dose). G1, G2 and G3 denote severity grades mild, moderate, and severe, respectively. See Methods for details regarding collection timepoints and severity grading. Vac = PfSPZ-LARC2 Vaccine; NS = normal saline placebo; N = sample size

| Number of participants with at least one: | All Vac<br>N=50 | All NS<br>N=25 | Group 1 (20-50 yr) |  | Group 2 (20-50 yr) |  | Group 3 (11-19 Yr) |  | Group 4 (6-10 yr) |  | Group 5 (1-5 yr) |  |
| --- | --- | --- | --- | --- | --- | --- | --- | --- | --- | --- | --- | --- |
|  |  |  | Vac: 2x10<sup>5</sup><br>N=10 | NS<br>N=5 | Vac: 4x10<sup>5</sup><br>N=10 | NS<br>N=5 | Vac: 2x10<sup>5</sup><br>N=10 | NS<br>N=5 | Vac: 1x10<sup>5</sup><br>N=10 | NS<br>N=5 | Vac: 1x10<sup>5</sup><br>N=10 | NS<br>N=5 |
| **Global Summary** |  |  |  |  |  |  |  |  |  |  |  |  |
| Solicited local | 0 | 0 | - | - | - | - | - | - | - | - | - | - |
| Solicited systemic | 9 (18%) | 3 (12%) | 5 (50%) | 1 (20%) | 1 (10%) | 2 (40%) | 2 (20%) | - | 1 (10%) | - | - | - |
| - grade 2 | 3 (6%) | 2 (8%) | 2 (20%) | 1 (20%) | 1 (10%) | 1 (20%) | - | - | - | - | - | - |
| - grade 3 | 0 | 0 | - | - | - | - | - | - | - | - | - | - |
| Unsolicited AE | 10 (20%) | 3 (12%) | 4 (40%) | 1 (20%) | 2 (20%) | 1 (20%) | 1 (10%) | - | 1 (10%) | - | 2 (20%) | 1 (20%) |
| - grade 2 | 4 (8%) | 1 (4%) | 2 (20%) | - | 1 (10%) | 1 (20%) | 1 (10%) | - | - | - | - | - |
| - grade 3 | 0 | 0 | - | - | - | - | - | - | - | - | - | - |
| Lab abnormality | 16 (32%) | 14 (56%) | 5 (50%) | 3 (60%) | 2 (20%) | 4 (80%) | 3 (30%) | 4 (80%) | 5 (50%) | 1 (20%) | 1 (10%) | 2 (40%) |
| - grade 2 | 7 (14%) | 3 (12%) | 2 (20%) | - | - | - | 3 (30%) | 1 (20%) | 2 (20%) | 1 (20%) | - | 1 (20%) |
| - grade 3 | 3 (6%) | 1 (4%) | - | - | - | 1 (20%) | - | - | 3 (30%) | - | - | - |
| Serious AE | 0 | 0 | - | - | - | - | - | - | - | - | - | - |
| **Solicited AEs (recorded for 28 days following vaccination)** |  |  |  |  |  |  |  |  |  |  |  |  |
| Chills | 0 | 1 (4%) | - | -- | - | 1 (G1) | - | - | - | - | - | - |
| Cough | 2 (4%) | 1 (4%) | 2 (G1)<sup>V3</sup> | - | - | 1 (G1) | - | - | - | - | - | - |
| Dizziness | 2 (4%) | 1 (4%) | 1 (G1)<sup>V1</sup><br>1 (G2)<sup>V2</sup> | - | - | 1 (G1) | - | - | - | - | - | - |
| Fatigue | 4 (8%) | 2 (8%) | 2 (G1)<sup>V1/3</sup><br>1 (G2)<sup>V2</sup> | 1 (G2)<sup>V2</sup> | 1 (G2) | 1 (G2) | - | - | - | - | - | - |
| Fever | 2 (4%) |  | 2 (G1)<sup>V3/3</sup> | - | - | - | - | - | - | - | - | - |
| Headache | 6 (12%) | 0 | 3 (G1)<sup>V2/3/3</sup><br>1 (G2)<sup>V3</sup> | - | 1 (G2) | - | 1 (G1) | - | - | - | - | - |
| Rash | 3 (6%) | 0 | - | - | - | - | 2 (G1) | - | 1 (G1) | - | - | - |
| Myalgia | 1 (2%) | 0 | - | - | 1 (G1) | - | - | - | - | - | - | - |
| **Unsolicited AEs (recorded for 28 days following vaccination)** |  |  |  |  |  |  |  |  |  |  |  |  |
| Conjunctivitis | 1 (2%) | 0 | - | - | 1 (G2) | - | - | - | - | - | - | - |
| Abdominal pain | 2 (4%) | 0 | 1 (G1)<br>1 (G2) | - | - | - | - | - | - | - | - | - |
| Dental caries | 1 (2%) | 0 | 1 (G1) | - | - | - | - | - | - | - | - | - |
| Gastroesoph. reflux | 0 | 1 (4%) | - | 1 (G1) | - | - | - | - | - | - | - | - |
| Oral candidiasis | 1 (2%) | 0 | - | - | - | - | - | - | - | - | 1 (G1) | - |
| Cut. larva migrans | 1 (2%) | 0 | - | - | - | - | - | - | - | - | 1 (G1) | - |
| Facial Injury | 1 (2%) | 0 | - | - | - | - | - | - | 1 (G1) | - | - | - |
| Skin abrasion | 1 (2%) | 0 | - | - | 1 (G1) | - | - | - | - | - | - | - |
| Back pain | 1 (2%) | 0 | 1 (G1) | - | - | - | - | - | - | - | - | - |
| Torticollis | 0 | 1 (4%) | - | - | - | 1 (G2) | - | - | - | - | - | - |
| Oropharyngeal pain | 1 (2%) | 0 | - | - | 1 (G2) | - | - | - | - | - | - | - |
| Rhinitis | 0 | 1 (4%) | - | - | - | - | - | - | - | - | - | 1 (G1) |
| Furuncle | 1 (2%) | 0 | - | - | - | - | 1 (G2) | - | - | - | - | - |
| Pruritus | 1 (2%) | 0 | 1 (G1) | - | - | - | - | - | - | - | - | - |
| **Laboratory abnormalities 7 days post vaccination** |  |  |  |  |  |  |  |  |  |  |  |  |
| ↓ hemoglobin | 1 (2%) | 0 | 1 (G1)<sup>V1</sup> | - | - | - | - | - | - | - | - | - |
| ↑ hemoglobin | 1 (2%) | 0 | - | - | 1 (G1) | - | - | - | - | - | - | - |
| ↓ lymphocyte | 0 | 0 | - | - | - | - | - | - | - | - | - | - |
| ↑ lymphocyte | 2 (4%) | 1 (4%) | - | - | - | - | 2 (G2) | - | - | 1 (G1) | - | - |
| ↓ neutrophil | 1 (2%) | 0 | 1 (G2)<sup>V2</sup> | - | - | - | - | - | - | - | - | - |
| ↑ neutrophil | 2 (4%) | 1 (4%) | 2 (G1)<sup>V2/3</sup> | - | - | 1 (G1) | - | - | - | - | - | - |
| ↓ platelet | 1 (2%) | 0 | - | - | - | - | 1 (G2) | - | - | - | - | - |
| ↑ platelet | 2 (4%) | 2 (8%) | 1 (G1)<sup>V1</sup> | - | - | - | - | 1 (G1) | 1 (G1) | - | - | 1 (G1) |
| ↓ leukocyte | 0 | 3 (12%) | - | 1 (G1)<sup>V1</sup> | - | - | - | 2 (G1) | - | - | - | - |
| ↑ leukocyte | 1 (2%) | 0 | 1 (G1)<sup>V2</sup> | - | - | - | - | - | - | - | - | - |
| ↑ AST | 0 | 0 | - | - | - | - | - | - | - | - | - | - |
| ↑ creatinine | 0 | 0 | - | - | - | - | - | - | - | - | - | - |
| Any abnormality D7 | 7 (14%) | 7 (25%) |  |  |  |  |  |  |  |  |  |  |
| **Laboratory abnormalities 28 days post vaccination** |  |  |  |  |  |  |  |  |  |  |  |  |
| ↓ hemoglobin | 2 (4%) | 0 | 1 (G1)<sup>V1</sup> | - | - | - | - | - | 1 (G3) | - | - | - |
| ↑ hemoglobin | 1 (2%) | 1 (4%) | 1 (G1)<sup>V3</sup> | - | 1 (G1) | - | - | - | - | - | - | - |
| ↓ lymphocyte | 0 | 0 | - | - | - | - | - | - | - | - | - | - |
| ↑ lymphocyte | 2 (4%) | 1 (4%) | - | - | - | - | 2 (G2) | - | - | 1 (G2) | - | - |
| ↓ neutrophil | 7 (14%) | 1 (4%) | 5 (G1)<sup>V1/1/2/3/3</sup><br>2 (G2)<sup>(V1/2)</sup> | - | - | 1 (G1) | - | - | - | - | - | - |
| ↑ neutrophil | 4 (8%) | 1 (4%) | 4 (G1)<sup>V1/1/2/3</sup> | - | - | 1 (G1) | - | - | - | - | - | - |
| ↓ platelet | 1 (2%) | 1 (4%) | - | - | - | - | 1 (G2) | - | - | - | - | 1 (G2) |
| ↑ platelet | 4 (8%) | 2 (8%) | 1 (G1)<sup>V1</sup> | - | - | - | - | 1 (G1) | 2 (G1) | - | 1 (G1) | 1 (G1) |
| ↓ leukocyte | 0 | 5 (20%) | - | 3 (G1)<sup>V1/2/3</sup> | - | - | - | 2 (G1) | - | - | - | - |
| ↑ leukocyte | 1 (2%) | 0 | 1 (G1)<sup>V2</sup> | - | - | - | - | - | - | - | - | - |
| ↑ AST | 0 | 1 (4%) | - | - | - | 1 (G1) | - | - | - | - | - | - |
| ↑ creatinine | 1 (2%) | 1 (4%) | - | - | 1 (G1) | 1 (G1) | - | - | - | - | - | - |
| Any abnormality D28 | 16 (32%) | 11 (44%) |  |  |  |  |  |  |  |  |  |  |

Unsolicited AEs occurred in 10/50 (20%) vaccine recipients and 3/25 (12%) placebo recipients (p=0.52) (**Table 1**), and were similarly mild or moderate. All were transient and deemed unrelated to vaccination.

LAs were common in both vaccine and placebo recipients but were predominantly mild and not clinically significant. All LAs resolved. Overall, LAs occurred in 32% of vaccine recipients and 56% of placebo recipients (p=0.08), with no pattern suggesting vaccine-related toxicity. One grade 3 laboratory abnormality (hemoglobin decrease) was observed in a group 4 vaccine recipient but was not considered clinically significant because of minimal change from baseline.

#### Serious adverse events

No serious adverse events (SAE) occurred after vaccination. One preimmunization hospitalization of a male in his fourth decade for suspected appendicitis resulted in exclusion from dosing.

#### Long-term safety follow-up

Follow-up at 12 and 24 weeks was completed by 96% and 93% of participants, respectively. No patterns of illness attributable to vaccination were identified, and no delayed safety signals were observed. Details are provided in the **supplementary material**. Cross-sectional malaria prevalence surveys by TBS were performed following approval of a protocol amendment at the 12-week follow-up in group 5 and the 24-week follow-ups in groups 1, 3, 4 and 5 (**table S2**). There were no differences in the frequency of positive results between vaccine and placebo groups. In addition, 11 clinical malaria episodes were diagnosed during the period extending from 28 days post vaccination to the 24-week follow-up, 7 cases in vaccinees and 4 cases in controls (**table S3**) and resolved with standard DHA-P treatment.

### Immunogenicity

IgG antibodies to the Pf circumsporozoite protein (CSP) were assessed by enzyme-linked immunosorbent assay (ELISA) at baseline and at 2, 4, and 12 weeks after immunization. Baseline anti-PfCSP antibody levels varied widely and were inversely associated with age, with higher baseline levels in adults than children (Spearman’s r = −0.739, p < 0.0001) (**Table 2**). Post-immunization, anti-PfCSP responses increased over baseline in vaccine recipients, measured either as seroconversion rates (**Table 2**), net increases over baseline (**Fig. 4, tables S4, S6, Figs. S3, S4**), or fold increases over baseline (**Fig. 4, tables S5, S7, Figs. S3, S4**). Seroconversion occurred after single dosing in younger age groups but required repeated dosing in adults who received 2×10^5^ PfSPZ (**Table 2**). Setting aside group 2, which received 4×10^5^ PfSPZ, a 2- to 4-fold higher dose than any other group, seroconversion rates, net increases, and fold increases in IgG over baseline showed an age-dependent pattern, with stronger responses observed after one dose of 2×10^5^ PfSPZ in adolescents than in adults, and still stronger responses after half that dose, 1×10^5^ PfSPZ, in the two younger groups (**Fig 4**), with antibody levels in group 4 vaccinees significantly higher than in controls. In preschool children (group 5), the 60% seroconversion rate after a single 1×10^5^ PfSPZ dose (**Table 2**) and the nearly 5-fold increase in antibody levels from baseline (**Fig. 4**) were significantly higher than controls and the three oldest vaccine groups.

**Fig. 4.**
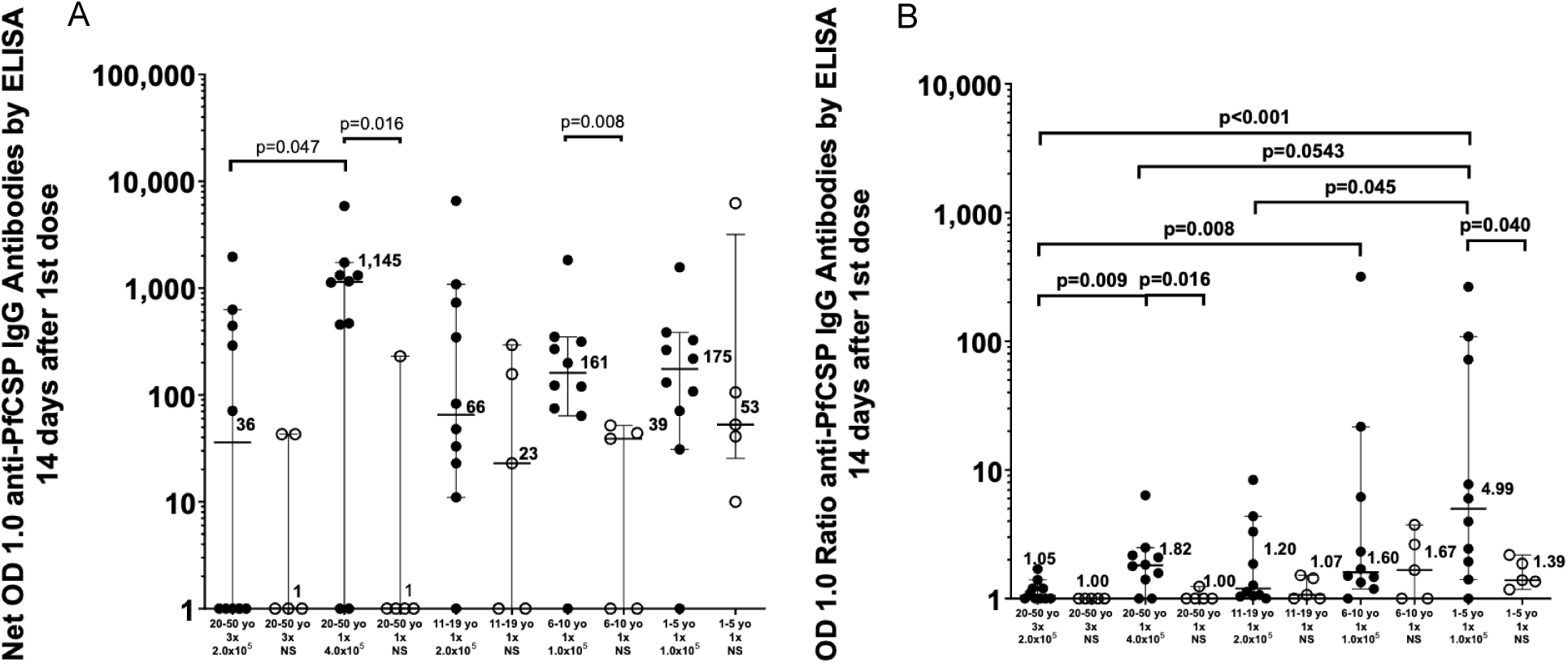
Age- and dose-dependent antibody responses to PfSPZ-LARC2 Vaccine. <u>Panel A</u>: Net IgG OD 1.0 responses to PfCSP measured 14 days after vaccination across doses and age groups were higher in vaccinees than controls, were higher after a single dose of 4×10^5^ PfSPZ in adults than a single dose of 2×10^5^ PfSPZ in adults, and also appeared higher in younger participants than older. The latter can be seen by comparing net IgG OD 1.0 levels between adolescents and adults receiving 2×10^5^ PfSPZ and between adolescents receiving 2×10^5^ PfSPZ and school age and pre-school children receiving half that dose. <u>Panel B</u>: Despite receiving only 1×10^5^ PfSPZ, pre-school children showed the most robust fold-increases over baseline.

**Table 2.**
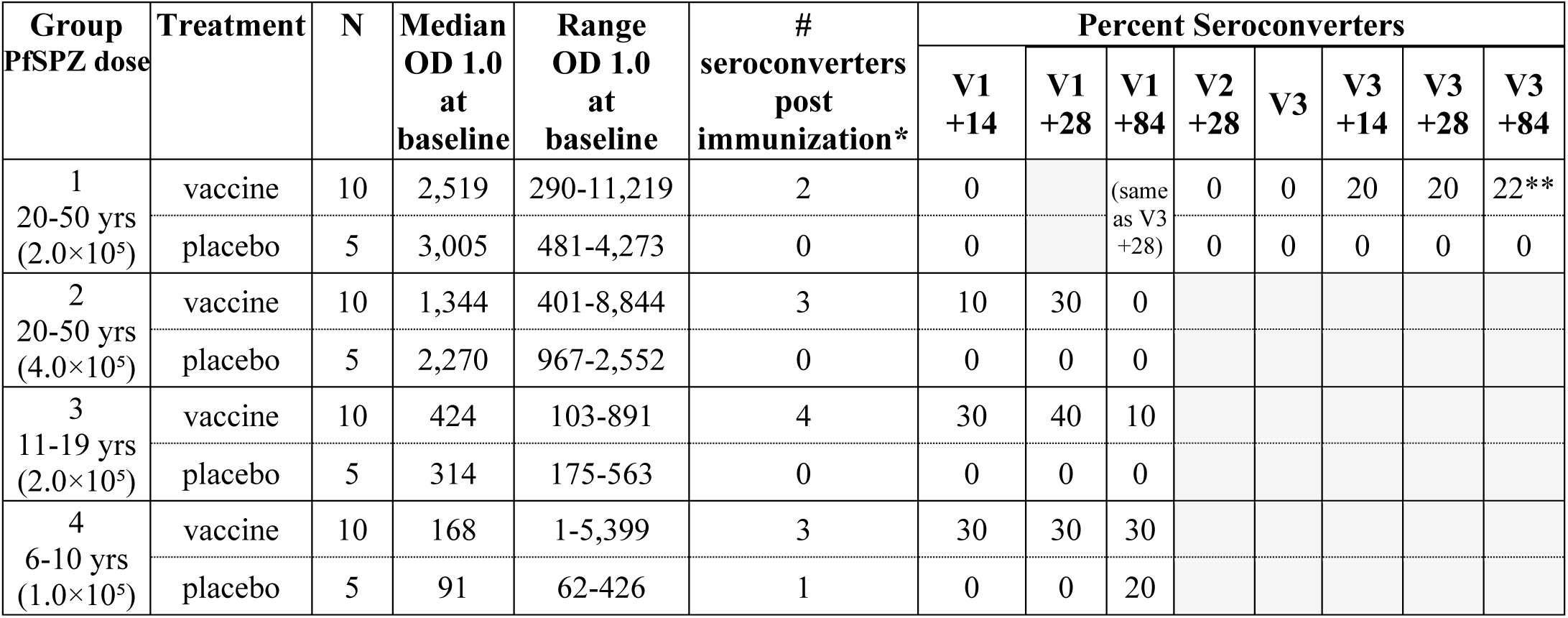
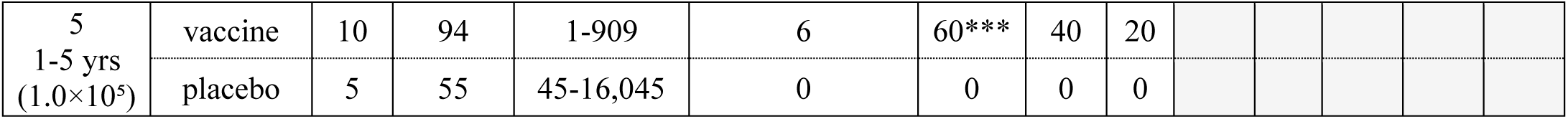
Baseline antibody levels and seroconversion rates. *Number of seroconverters at any timepoint. ** No sera available from one vaccinee. *** p=0.04 comparing group 5 vaccinees and placebos. Shading indicates no blood sample collected or time point not applicable.

In Group 1 adults receiving three doses of 2×10^5^ PfSPZ, antibody levels increased progressively after each dose with a significant higher overall level of antibodies in vaccinees as compared to controls at 4 weeks after the third dose when levels peaked (median net OD 1.0 = 2,749 for vaccinees and median net OD 1.0 =14 for controls) (**Fig. 5**) Documenting seroconverters required three doses (**Table 2**). In group 2, after a single dose, antibody responses to 4.0×10^5^ PfSPZ were significantly higher than after the first dose of 2×10^5^ PfSPZ in group 1, indicating a dose response (**Fig. 4**). IgM and IgA antibody levels showed no differences between vaccinees and controls, and no individuals in any treatment group met criteria for seroconversion (t**able S6**).

**Fig. 5.**
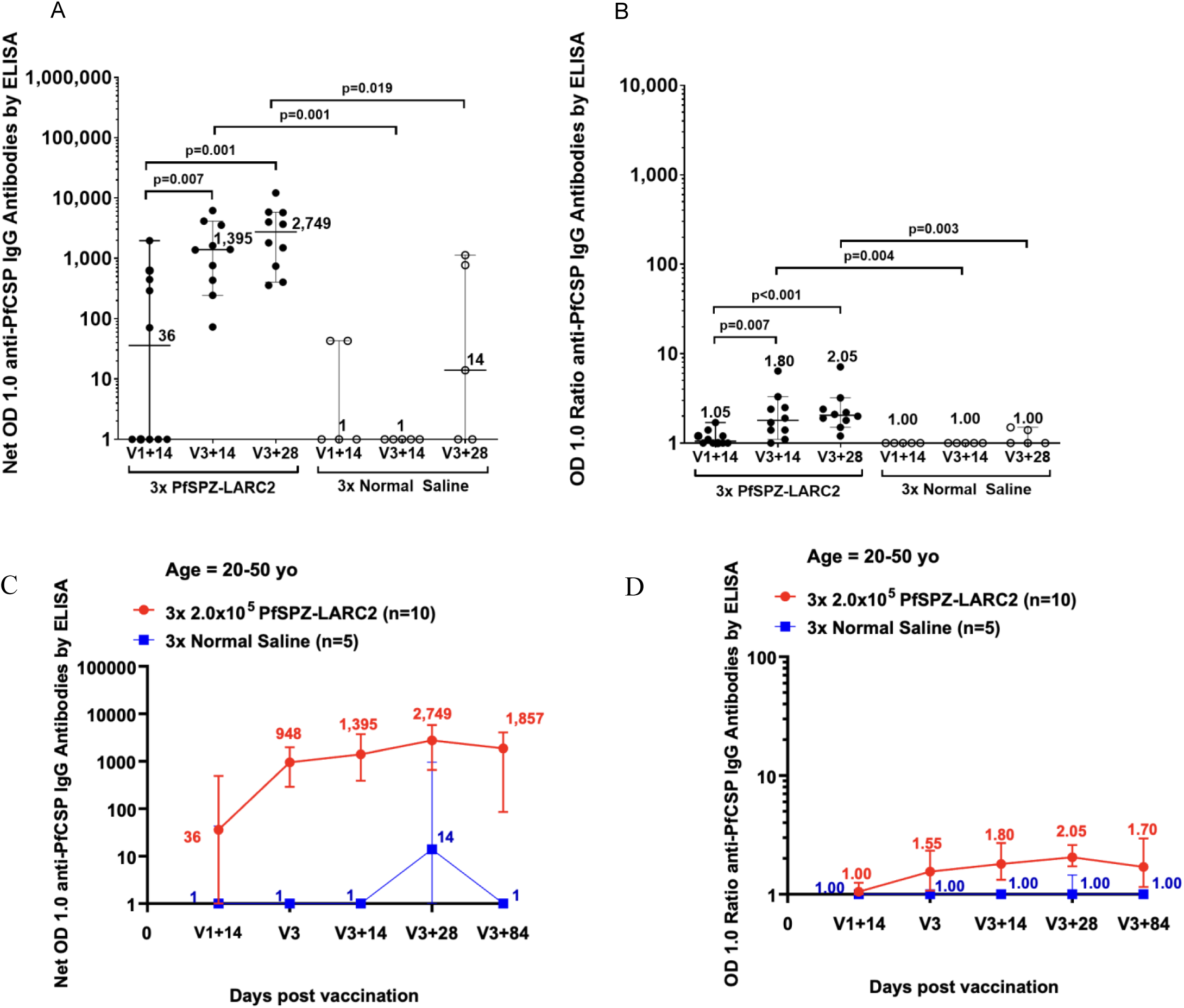
Antibody responses in adults receiving three doses of 2×10^5^ PfSPZ. Serum IgG responses to Pf circumsporozoite protein (PfCSP) measured by ELISA in adults receiving three doses. Antibody levels increased after repeated immunization compared to placebo.

## DISCUSSION

This first-in-human clinical trial of PfSPZ-LARC2 Vaccine in healthy 1- to 48-year-old residents of Sabou, Burkina Faso, demonstrated: 1) full attenuation (no breakthrough infections), 2) tolerability and safety (no statistically significant differences in AEs or laboratory abnormalities comparing recipients of vaccine and normal saline), and 3) immunogenicity (increased anti-PfCSP net OD 1.0 antibody levels and/or OD ratios post immunization) with the most robust seroconversion rate and relative increase of antibody responses in 1-5-year-olds.

The critical safety finding was the absence of breakthrough infections, indicating a successful translation of genetically altered phenotypes from rodent (*P. yoelii*) to human (Pf) malaria. Groups 1 to 4 were immunized first, before the peak rainy season, and no participants developed blood-stage parasitemia by thick blood smear during the 28 days of follow-up. Immunization of 1-5-year-olds followed, but only after local malaria transmission was underway. Three of 10 vaccine recipients developed Pf malaria (33%), one developed *P. ovale* malaria (10%), and two of five controls developed Pf malaria (40%). However, no infecting parasites had the *Mei2* deletion on molecular analysis, indicating that all were naturally transmitted wild-type infections. In summary, we detected no breakthrough Pf infections in any study group.

To counter the potential inhibitory effect on PfSPZ infection of naturally acquired immunity (NAI) and perform a stringent assessment of safety, we administered PfSPZ-LARC2 Vaccine at 62.5 to 125 times the non-immune adult 100% infectious dose of PfSPZ Challenge to adolescents and adults and 31.25 times the non-immune adult 100% infectious dose to 1-10-year-olds (*23*). We expected that this high-dose inoculum would overcome NAI, and, if attenuation were incomplete, result in detectable blood-stage parasitemia in at least a subset of participants. Because there were no blood-stage infections, our results indicate that LARC2 parasites arrest development in the late liver stage as shown in humanized mice and that this vaccine is safe to proceed to larger studies in Africans and also to test for attenuation in non-immunes. Both types of studies are ongoing.

Naturally transmitted Pf sporozoites do not cause clinical symptoms, signs, or pathology when they are in the bloodstream on the way to the liver or during the 5-6 days they develop as liver stages within hepatocytes. Clinical manifestations occur only after the asexual parasites released from hepatocytes have multiplied in the blood through several cycles and exceed densities of 1 to 100 parasites/μL. These characteristics of human malaria led us to anticipate that PfSPZ vaccines, which arrest development during the liver stages, would be equally well tolerated as naturally transmitted sporozoites. This expectation has been largely supported by the many trials of PfSPZ vaccines. For the early arresting, non-replicating PfSPZ Vaccine, PfSPZ-GA1, and PfSPZ-CVac (pyrimethamine), there has been no evidence of side effects (*9–15, 24–32*) except an excess of cases of myalgia in one study (*13*) and of subjective fever in another (*32*). For the fully replicating PfSPZ-CVac (chloroquine), there have been no differences between vaccinees and controls during the first 6 days after inoculation, which is before the development of blood-stage parasitemia and associated symptoms (*8, 11, 18, 33*). The findings of this trial – adverse event profiles indistinguishable between vaccinees and controls – are consistent with this experience, and reinforce the conclusion that LARC2 parasite development arrests in the liver.

PfSPZ-LARC2 Vaccine was immunogenic. The IgG PfCSP antibody analysis showed a dose-response in adults. At 2 weeks after a single dose of 2×10^5^ PfSPZ, the median net OD 1.0 was 36, while after a single dose of 4×10^5^ PfSPZ, the median net OD 1.0 was 1,145 (p = 0.047) (**Fig. 4**). The peak median net OD 1.0 after 3 doses of 2×10^5^ PfSPZ was 2,749 at 28 days after the third dose as compared to a median net OD 1.0 of 14 in the normal saline controls at this time point (p = 0.019). Despite the significantly higher level of antibodies as compared to the controls after the third dose, the rigorous criteria identified a low rate of seroconversion of 20% in the adults who received 3 doses of 2×10^5^ PfSPZ (**Table 2**). The rates of seroconversion after a single dose of PfSPZ increased with decreasing age and was highest in the 1-5-year-olds (60% after 1 dose of 1×10^5^ PfSPZ). The low adult seroconversion rate in group 1 after 3 doses was similar to the 28% seroconversion rate in Malian adults after 3 doses of 2.048×10^5^ PfSPZ of PfSPZ-CVac (chloroquine) (**fig. S6**) (*34*). PfSPZ-CVac (chloroquine) has never been given to children. However, in Tanzania (*25*) and Equatorial Guinea (*30*), 1-5 year olds immunized with PfSPZ Vaccine had better antibody responses to PfCSP than did adults immunized with the same dosage regimen. The finding of increased antibody responsiveness with younger age in this and prior studies supports the hypothesis that cumulative exposure to malaria with increasing age leads to a reduced ability to develop antibody responses to Pf antigens such as PfCSP, a condition sometimes called immunotolerance. Intrinsic differences in the responsiveness of the immune system related to age may also play a role, as could the PfSPZ administered per weight or body surface area.

The next step in clinical development of PfSPZ-LARC2 Vaccine will be to assess efficacy, which we expect to be as good as PfSPZ-CVac (chloroquine), which was 8 times more potent than the early-arresting replication-deficient PfSPZ Vaccine in German adults (*12, 18*). PfSPZ-LARC2 Vaccine has the potential to be even more protective than PfSPZ-CVac (chloroquine) for several reasons: 1) Asexual blood-stage parasitemia is present on days 7-9 after doses of PfSPZ-CVac (chloroquine), and asexual blood-stage parasitemia suppresses cellular immune responses to malaria liver stages (*35, 36*). Since there appears to be no blood-stage parasitemia with PfSPZ-LARC2 Vaccine, immune responses are expected to be more robust. 2) Chloroquine is an immunosuppressant. Since chloroquine is not used with PfSPZ-LARC2 Vaccine, immune responses may again be more robust. 3) With PfSPZ-CVac (chloroquine), Pf parasites are released into the bloodstream at the end of the liver stage in packets called merosomes. These circulate to other parts of the body, especially the lungs, rupture, and release merozoites, which invade erythrocytes. In contrast, the parasite material from disintegrating PfSPZ-LARC2 liver stage parasites is released locally in the liver and carried to the lymph nodes that drain the liver. This should facilitate local induction of immune responses compared to those induced in the lung, enhancing liver-resident cellular immunity.

The hypothesis regarding increased potency of PfSPZ-LARC2 Vaccine as compared to PfSPZ-CVac (chloroquine) is supported by clinical data with a parasite called GA2 or PfSPZ-LARC1. As described earlier, this parasite was generated by a team at Seattle Children’s Research Institute by deleting the *mei2* gene (LARC1), one of the two genes deleted in PfSPZ-LARC2 (*20, 21*). LARC1 was later independently recreated at Leiden University Medical Center as GA2 by deleting the *mei2* gene (*20*). When 10 non-immune adults were immunized by a single exposure to 50 GA2 infected mosquitoes, there were no breakthrough infections and 9 of 10 (90%) recipients were protected against homologous CHMI 6 weeks after immunization (*37*). This high degree of efficacy after a single immunization is unprecedented. Of note, 3 sessions of 50 infected mosquitoes gave the same level of protective efficacy as one session (*38*).

The study reported here had limitations. The sample size was modest, and protective efficacy was not assessed, which restricts interpretation of immunogenicity in relation to potential efficacy. Although no vaccine strain breakthrough infections were observed, larger studies, particularly in malaria-naive individuals, are required to confirm the robustness of attenuation. In addition, the durability of immune responses is not yet established. Other studies are evaluating efficacy, exploring optimized dosing regimens, and further characterizing immune responses.

In conclusion, PfSPZ-LARC2 Vaccine administered at 2×10^5^ or 4×10^5^ PfSPZ in malaria-exposed adults, 2×10^5^ PfSPZ in 11–19-year-olds, and 1×10^5^ PfSPZ in 6-10- and 1-5-year-olds demonstrated safety and tolerability, and showed no evidence of breakthrough blood-stage infection, indicating successful attenuation of the vaccine parasite. It induced antibodies to PfCSP with the most robust responses in younger age groups. These results provide the foundation for an ongoing Phase 2 clinical trial now underway in Burkina Faso aiming for the eventual licensure and implementation of the world’s first genetically attenuated vaccine against a disease caused by parasites.

## MATERIAL and METHODS

### STUDY DESIGN

#### Research Objectives

BFSPZL1 was a single-center, first-in-humans, Phase 1 trial to investigate the safety, tolerability, and immunogenicity of PfSPZ-LARC2 Vaccine. Because the parasite is genetically engineered to halt parasite development after replication in the liver, surveillance for breakthrough blood-stage infection (parasitemia) was a focus of the safety evaluation as incomplete attenuation would preclude further testing and development.

#### Research participants

The trial was conducted in healthy, malaria-exposed adults, adolescents, school children, and pre-school children living in Sabou, Burkina Faso. Participation from pre-immunization drug clearance to last study visit lasted 27 (groups 2-5) to 35 (group 1) weeks.

#### Experimental design

The trial was randomized, double-blind, and placebo-controlled. Normal saline was selected as the control because it should cause no side effects, is indistinguishable from the vaccine once loaded into a syringe, and can be safely administered by direct venous inoculation (DVI)^1^. The vaccine was tested as shown in **Fig. 2**. There were no significant changes to the protocol after the trial began. There was no public or participant involvement in the design of the trial, which was registered at clinicaltrials.gov (NCT06652737). The trial is complete.

#### Oversight

Ethical approval was obtained from the Comité d’Ethique pour la Recherche en Santé (CERS) in Burkina Faso and the Institutional Review Board of the University of Maryland, Baltimore. Regulatory oversight was provided by Agence Nationale de Régulation Pharmaceutique (ANRP) in Burkina Faso and the US Food and Drug Administration (FDA). Sanaria Inc. sponsored the trial under a US FDA investigational new drug (IND) application.

#### Study team and site

The Groupe de Recherche Action en Santé (GRAS) conducted the trial in the Sabou Health District (SHD) about 100 kms west of Ouagadougou, the capital of Burkina Faso. The SHD covers an area of 449 km^2^ in the region of Boulkiemde with 112,485 inhabitants. Most people living in small rural villages in houses made with mud or cement walls and thatched or metal roofs. The population is stable and mainly composed of the Mossi ethnic group, with farming the primary activity. The study’s field station, the Antenne de Recherche Clinique de Sabou, is located inside the Saint Maximilien Kolbe Hospital in Sabou town. The SHD includes 20 peripheral health facilities serving as the primary points of contact with the health system. Sabou has a Sahelian eco-climatic zone with a single rainy season from June to October characterized by intense malaria transmission followed by a long dry season during which there is very limited malaria transmission. Pf is the primary malaria parasite in the region with *P. malariae* and *P. ovale* occasionally seen (only *P. ovale* was identified during this trial).

#### Investigational product (IP)

PfSPZ-LARC Vaccine is composed of aseptic, purified, cryopreserved, live (metabolically active) PfSPZ diluted post thaw to 0.5 mL in phosphate buffered saline and 1% human serum albumin. The PfSPZ are genetically attenuated by the deletion of *Mei2* and *LINUP* genes. *Mei2^-^/linup^-^* parasites replicate in the liver and grow to a size comparable to normal liver stage schizonts but are unable to transition to blood-stage parasites in the FRG humanized mouse model and disintegrate spontaneously starting about day 7 (*21*). PfSPZ products are manufactured in aseptic mosquitoes by Sanaria Inc. (Rockville MD), purified and stabilized in liquid nitrogen vapor phase at −150 °C to −196 °C. The placebo was sterile, non-pyrogenic 0.9% sodium chloride for injection acquired commercially (Hospira (Pfizer)). *Stopping rules:* The SMC was empowered to stop study progression to the next cohort in case of breakthrough or concerning events, including a serious AE deemed possibly related to vaccination.

#### Study procedures

##### Screening

Participants were screened to ascertain good health. Enrollment criteria included normal electrocardiogram, no history of seizures other than simple febrile seizures, no history of migraine headaches, sickle cell disease, chronic illness or poor tolerance to antimalarial drugs, absence of hepatitis B, hepatitis C, or HIV infection, and no recent or planned use of drugs with antimalarial activity or other vaccines. Women of child-bearing potential were required to use highly effective birth control.

##### Randomization and masking

Randomization assigned participants 2:1 to vaccine or normal saline placebo within each group, each pilot subgroup, and each main subgroup. Treatment assignment tables were generated by an independent biostatistician (TASK Clinical, Cape Town, South Africa) using a randomly permuted block design and provided to the unblinded GRAS pharmaceutical operations staff. Syringes were filled in a locked pharmacy and passed to the clinical team labeled with the participant ID. Diluted vaccine and normal saline are clear, non-viscous fluids that are indistinguishable. Pharmacy staff did not interact with trial participants. Blinding of the GRAS and sponsor clinical teams, the external monitors (ClinaPharm, Cotonou, Benin), clinical and research laboratories and SMC was maintained until data were cleaned and locked.

##### Drug clearance prior to immunization

All participants received a three-day treatment course of artemether/lumefantrine combination tablets (AL) (80/480 mg twice per day) two weeks before immunization to clear any parasitemias, which suppress PfSPZ-induced immunity even when densities are below the threshold of detection by thick blood smear (*15*).

##### Investigational product administrations

IP was administered by direct venous inoculation (DVI), injecting the 0.5 mL syringe contents over 1 or 2 seconds through a 25-gauge needle inserted in a superficial arm vein after demonstrating blood flashback. Injection had to be completed within 30 minutes of PfSPZ thaw. DVI is nearly painless as no tissue planes are disrupted by the injection. DVI allows efficient delivery of PfSPZ to the liver sinusoids, where PfSPZ marginate and invade hepatocytes.

##### Safety follow-up post immunization

Participants were seen on days 1 *or* 2 and 3 *or* 4 post-immunization for recording of AEs possibly resulting from PfSPZ or liver stage parasites. Starting on day 6, when AEs could result from breakthrough blood-stage parasites, participants were seen daily (adult groups) or every other day (pediatric groups) through day 20 or 21, then every two to three days until day +8 (**Fig. 2**). At each visit, a clinical evaluation was performed, and from day 6 onwards, thick blood smears were made and read the same day (**Fig. 2**). Participants (or their parents) were contacted by telephone or visited for additional AE assessments on non-thick blood smear days between day 6 and 28. Thus, there was contact between the clinical team and the participant for 23 consecutive days, from day 6 to 28 after immunization (or first immunization in Group 1). In Group 1, visits were on days 7, 14, 21 after second and third immunizations. Medical consultation was available 24/7 to all participants through 24 weeks post-immunization. Laboratory tests (absolute leukocyte count, hemoglobin, platelets, alanine aminotransferase, and creatinine) were performed on days 6 and 28 after each immunization.

##### Drug clearance post immunization

All participants received a full three-day treatment course of dihydroartemisinin/piperaquine combination tablets (DHA-PP) (160/1,280 mg daily) to eliminate any undetected parasitemia starting on day 28 after immunization (after the last immunization for group 1). The same regimen was used for any malaria infections diagnosed by thick blood smear during the trial. Six participants in Group 5 received terminal treatment with artesunate-pyronaridine rather than DHA-PP because they had recently received DHA-PP to treat wild-type infections acquired during the immunization period (back-to-back courses of DHA-PP are not recommended due to potential prolongation of QTc interval).

##### Thick blood smears

These were prepared by pipetting 10 μl of fresh blood onto a 1 x 2 cm rectangular template on a glass slide and allowing the blood to dry, followed by Giemsa staining. The microscopist read 2 μl of blood with viewing of a single unequivocal parasite as positive, yielding a 0.5 parasites/μl limit of detection.

##### Antibody assays

IgG, IgM, and IgA antibodies to Pf circumsporozoite protein [PfCSP] were assessed by ELISA in sera taken before and after immunization using published methods (*8*).

##### Genetic identity test

During the 28 days after administration of PfSPZ-LARC2 Vaccine or placebo, if malaria parasites were identified by TBS, participants were treated with DHA-PP. Before treatment, blood samples were taken and placed on filter paper as dried blood spots (DBSs). A PCR-based assay was used to differentiate *mei2^-^*/*linup^-^*parasites from wild-type parasites (**Fig. 3**).

###### Outcomes

The pre-specified primary safety outcomes were (1) the number of trial participants with blood-stage infection during the first 28 days after immunization shown to be the *mei2^-^*/*linup^-^*vaccine strain of Pf; (2) the incidence of (a) grade 3 solicited AEs in the 28 days post immunization; (b) related grade 3 unsolicited AEs in the 28 days post immunization; (c) grade 3 abnormal laboratory values one week after immunization; (d) related serious adverse events (SAEs) throughout the study period. The safety population consisted of all participants receiving any investigational product.

### STATISTICAL ANALYSIS

#### Sample size

Target sample size of 15/group (with 3 serving as pilots) was based on safety considerations in this first-in-humans trial. The safety objective was to demonstrate, with 90% confidence, that the true rate of break-through infections was less than 5%. After the first cohort (which included 20 vaccine recipients) completed 28 days of follow-up, as there were no breakthroughs, there was 90% confidence that the true rate of breakthrough was less than 12%. This was deemed sufficient to proceed to the two older pediatric groups, which also included 20 vaccine recipients. After their 28 days of follow-up, as there were no breakthroughs, there was 90% confidence that the true rate of breakthrough was less than 6%. This was deemed sufficient to proceed to the youngest age group. As there were no breakthroughs after completing follow-up in the third cohort, which included 10 vaccine recipients, the study achieved 90% confidence that the true rate of breakthrough was less than 4.6%, addressing this primary safety objective.

#### Statistical tests

Comparisons of the number of participants experiencing AEs and seroconversion rates were analyzed using two-tailed Fisher’s exact test. Comparisons of antibody levels were analyzed using a two-sided Mann-Whitney test and non-parametric analysis of variance. The statistical software used was SAS^®^ version 9.4 or later (SAS Institute Inc., Cary, NC, USA) or R software version 4.3.1 or later. No corrections were made for multiple comparisons.

## List of Supplementary Materials

Table S1-S6

Figures S1-S6

## Data Availability

Data and study documents are available on reasonable request to the corresponding authors.

## Acknowledgements

We gratefully acknowledge the support of the Safety Monitoring Committee: Fla Koueta, who also served as the local safety monitor, Andrea Berry (committee chair) and Joel Goldberg. We additionally thank the following team members: GRAS – Lydia M. Dabre, clinical trial assistant; UMB – Melissa Myers, Fleesie Hubbard; Sanaria – Anusha Gunasekera, Jeremy Guth, Dimitri Koutsoumis, Maria Orozco, Asha Patil, Shachi Shah, and the Sanaria manufacturing team; TASK – Johann de Bruyn, Kerissa Chetty, Joni Mostert, Boipelo Motsamai, Mojalefa Petje, Edwin Tshegofatso Mohlake; ClinaPharm – Florence Amoussou, Nawal Tigri, Ines Zevounou.

## Author contributions

T.L.R., S.L.H, A.O, A.B.T, S.B.S., M.L. and S.H.I.K conceived and designed the trial; S.H.I.K., N.C., G.D., and A.M.V. created the vaccine; S.B.S., A.O., A.B.T., B.I.S., A.R.Z., I.N, J.M.K., N.K., R.K.M., S.E., E.S.B., S.Y.O., N.H., J.B.Y. conducted the trial; T.L.R, S.L.H., B.K.L.S, T.M., M.C.C., N.K.C., Y.A., E.R.J. provided sponsor oversight and clinical, regulatory, pharmaceutical operations, and immunological support; N.K.C. performed the anti-PfCSP assays and analyses. T.L.R, S.L.H, T.M., and S.B.S. analyzed the results; T.L.R., S.L.H., M.L., S.H.I.K, and S.B.S. wrote the manuscript.

## Data and materials availability

Data and study documents are available on reasonable request to the corresponding authors.

