## Supplementail tables and figures for "First-in-human trial in Burkinabe adults and children of replication-competent, genetically attenuated PfSPZ-LARC2 Vaccine for malaria"

### <u>SUPPLEMENTAL MATERIAL</u>

<u>Participant Demographics</u>

Demographic data from the trial participants are provided in **Table S1**. Treatment assignment was reasonably well balanced with respect to age, height, weight and BMI. There were some imbalances in sex (e.g., group 4 had more females than males among vaccinees and the reverse for placebo recipients).

#### Table S1: Demographic Data

| Category | Measure | G1 (adult) | | G2 (adult) | | G3 (11-19 yr) | | G4 (6-10 yr) | | G5 (1-5 yr) | |
|---|---|---|---|---|---|---|---|---|---|---|---|
| | | Vac N=10 | NS N=5 | Vac N=10 | NS N=5 | Vac N=10 | NS N=5 | Vac N=10 | NS N=5 | Vac N=10 | NS N=5 |
| Age (years) | Mean | 39.7 | 38.0 | 28.0 | 36.2 | 14.6 | 15.0 | 6.7 | 7.2 | 4.0 | 3.0 |
| | Min/Max | 26-48 | 27-44 | 20-45 | 24-47 | 11-17 | 13-17 | 6-8 | 6-8 | 2.-5 | 1-4 |
| Sex | Male | 5 | 2 | 6 | 4 | 6 | 4 | 3 | 3 | 6 | 1 |
| | Female | 5 | 3 | 4 | 1 | 4 | 1 | 7 | 2 | 4 | 4 |
| Height (cm) | Mean | 168.8 | 166.4 | 167.1 | 174.8 | 158.2 | 154.6 | 124.7 | 124.0 | 101.8 | 98.6 |
| | Min/Max | 158-179 | 155-175 | 152-179 | 169-182 | 143-175 | 139-166 | 116-137 | 115-129 | 90-115 | 86-117 |
| BMI | Mean | 22.4 | 21.6 | 22.5 | 22.3 | 18.3 | 17.9 | 13.4 | 13.3 | 14.6 | 14.6 |
| | Min/Max | 19.0-26.3 | 18.3-26.3 | 18.1-25.7 | 18.1-26.1 | 15.3-21.5 | 15.7-20.3 | 12.4-15.6 | 12.2-14.9 | 13.5-15.8 | 12.3-15.8 |
| Weight (kg) | Mean | 64.2 | 60.0 | 62.9 | 68.3 | 46.1 | 43.0 | 20.9 | 20.5 | 15.3 | 14.5 |
| | Min/Max | 47.3-84.1 | 51.7-80.6 | 46.4-74.2 | 54.9-80.0 | 31.3-57.9 | 35.4-55.9 | 16.9-26.7 | 16.1-23.6 | 11.3-19.3 | 9.1-21.7 |

<u>Participant Flow and Reasons for Dropouts (details)</u>

74/75 (98.7%) enrolled (immunized) participants completed follow-up through presumptive treatment on days 28-30 after the last dose of vaccine or placebo (days 84-86 for group 1). One vaccine recipient in group 3p (pilot) (#99) left the country on day V+17 with a relative (#69), a vaccine recipient in group 2m (main) who had completed follow-up through presumptive treatment

(**Fig. 1**). The group 3p participant did not receive terminal treatment, but telephone follow-up established ongoing health without malaria infection. Neither participant returned for 12- or 24-week follow-ups.

There were four post-presumptive treatment losses in addition to #69: one group 1m vaccine recipient (#36) and one group 3m vaccine recipient (#110) missed 12 and 24 week follow-ups (left country and withdrew consent, respectively) and one group 2m placebo (#61) and one group 5m vaccine recipient (#134) missed the 24 week follow-up (lost to follow-up and left country with parents, respectively) (**Fig. 1**).

In summary, all immunizations were administered, 1/75 (1.3%) participants did not complete 28 days of follow-up and presumptive treatment, an additional 3/75 (4.0%) missed follow-ups at 12 and 24 weeks, and an additional 2/75 (2.7%) missed follow-up at 24 weeks.  Thus, 69/75 (92%) enrolled participants completed the study.

<u>Malaria Infections Identified in Group 5 During Initial 28 Days of Follow-up (details)</u>

All seven of the malaria infections identified in group 5 by thick blood smear (**Fig. 3A**) were treated with a 3-day course of DHA-P. Six of the seven were retrospectively confirmed as Pf (N=5) or *P. ovale* (N=1) infections at the University of Washington using a PCR-based genotyping identity assay (**Fig. 3C, D, E**). The infection in the seventh child was shown to be a false positive by the same assay and re-evaluation of the thick blood smear. None of the six confirmed infections was symptomatic and none had fever. Five of the six were clustered in a 6-day period during the first two weeks of group 5 follow-up, while one occurred during the second two weeks of follow-up, not a statistically significant difference (5/15 vs. 1/9 susceptible participants, p = 0.35, Fishers exact test). Because of treatment with DHA-P, five of the six were administered artesunate-pyronaridine for terminal treatment days 28-30 as a safety measure to avoid the additive QTc prolongation effects that could occur with back-to-back DHA-P treatments, while one received a second course of DHA-P (no QTc prolongation observed on ECG).

Key amplicons were sequenced.

(1) To further the analysis of the parasite isolated from participant 122, the lower amplicon band seen in the fourth lane of the gel depicted in **Fig. 3D** was sequenced and was shown to correspond to the *Plasmodium ovale* MEI2 gene.

**Figure S1: Sequencing Amplicon from Fig. 3D**

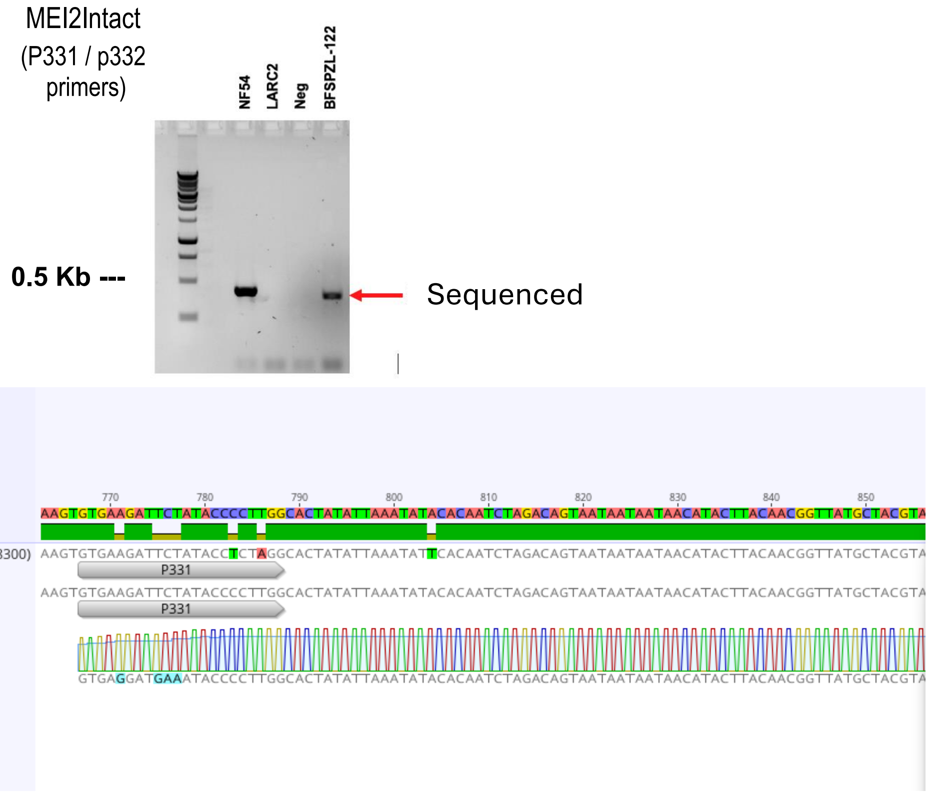


sample was extracted and analyzed by PCR. Fresh extracted material was used for genotyping PCR yielding the expected band and additional band slightly larger. The two bands were sent for sequencing, and the analysis confirmed the lower amplicon band corresponded to *P. falciparum MEI-2* sequence.

**Figure S2: Sequencing Amplicons from Fig. 3E**

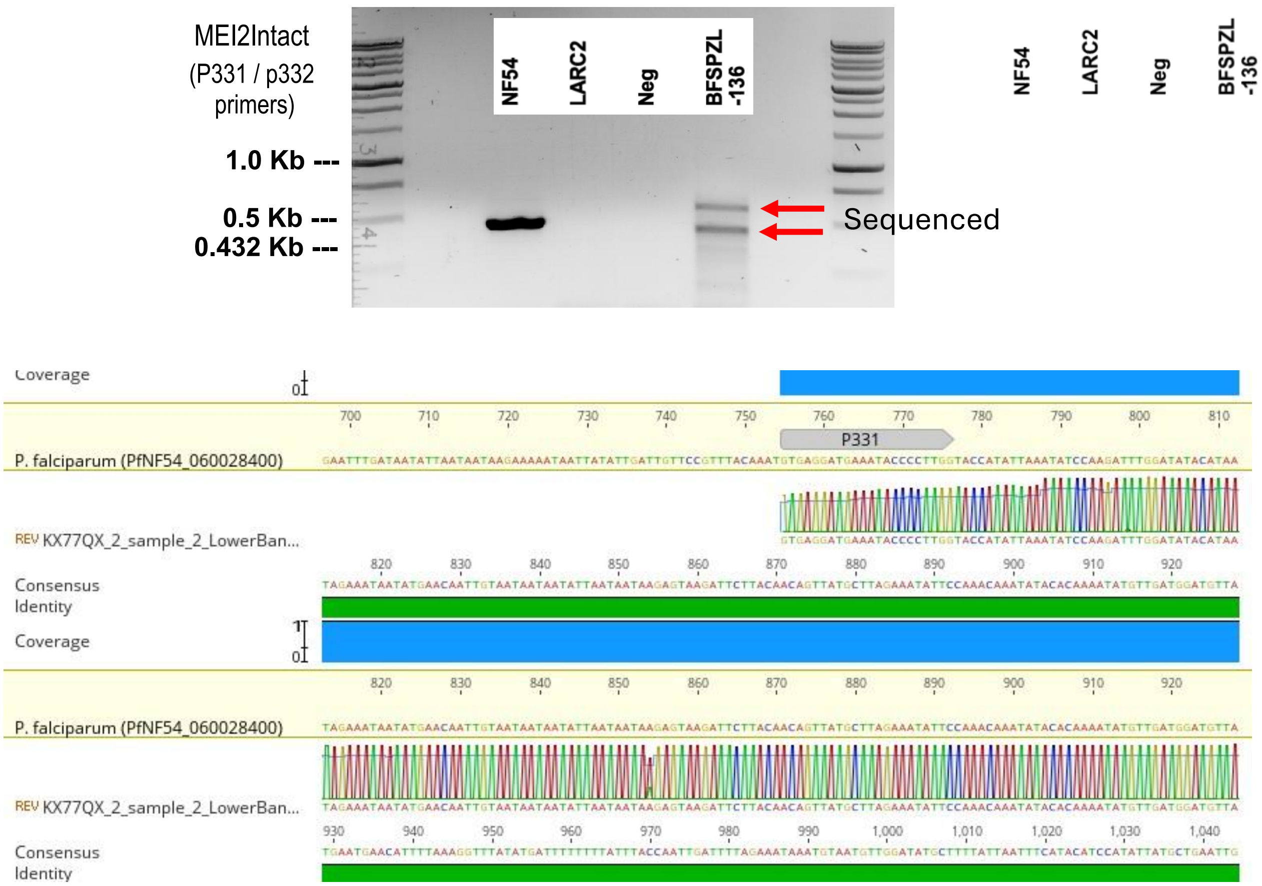


<u>Assessment of Adverse Events and Laboratory Abnormalities</u>

Solicited local (site of injection) adverse events (AEs), solicited systemic AEs, and unsolicited AEs were followed after each immunization for 7, 28 and 28 days, respectively. Laboratory testing was done at screening, pre-immunization, 6 or 7 days after immunization, and 28 days after immunization.

- *Solicited Adverse Events Severity Grading:*

  - Objective fever (axillary): Grade 1 (mild): 37.5°C to 38.4°C; Grade 2 (moderate): 38.5°C to 38.9°C; Grade 3 (severe): ≥39.0°C

- Local solicited AEs: erythema, swelling, induration, bruising: Grade 1: 0.5 – 5 cm; Grade 2: >5.1 – 10 cm; Grade 3: >10 cm, necrosis, exfoliative dermatitis. Findings <0.5 cm not recorded.

- For AEs other than objective fever and local solicited AEs, severity was graded by the effect on activities of daily living: mild (Grade 1): no restrictions; moderate (Grade 2): some restrictions; severe (Grade 3): interrupts activities of daily living (can't work, can't go to school).

- *Solicited Adverse Events Relationship to Immunization:* All solicited AEs were assumed to be related to immunization.

- *Unsolicited adverse events Severity Grading:* Mild, moderate or severe (same criteria as with solicited AEs).

- *Unsolicited Adverse Events Relationship to Immunization:* Unrelated, unlikely related (combined as "unrelated"), or possibly, probably or definitely related (combined as "related").

- *Laboratory Abnormalities Clinical significance:* Clinically significant or non-clinically significant.

- *Laboratory Abnormalities Severity:* Graded as mild, moderate, or severe by pre-determined thresholds.

- *Laboratory Abnormalities Relationship to Immunization:* this was not assessed.

Adverse Events Recorded

*Systemic local AEs:* Solicited local AEs were tenderness and induration to palpation, bruising, erythema and swelling by visualization, and pain and pruritus by questioning. No solicited local AEs >2.5 cm in diameter were recorded in any participants (**Table 1**). Solicited systemic AEs for all groups were fever, defined as axillary temperature > 37.5°C , rash, urticaria and edema by observation, and for groups 1-4, pruritus, subjective fever, headache, malaise, fatigue, chills, myalgia and arthralgia by questioning. Because group 5 participants were too young to question, the latter AEs were replaced by questioning the parents about drowsiness, irritability, and inability/refusal to eat or drink. All AEs were assessed for severity as mild/grade 1 (not interfering with daily activities), moderate/grade 2 (some interference) or severe/grade 3 (preventing daily activities).

*Systemic solicited AEs:* These were identified in 9/50 (18%) vaccine recipients and 3/25 (12%) placebo recipients (p=0.74) (**Table 1**). These were mild/grade 1 in severity except for 3 (6%) and 2 (8%) participants in vaccine and placebo groups, respectively, who experienced moderate/grade 2 AEs. All grade 2 AEs occurred in the adult groups: after the first immunization, headache and fatigue (accompanied by mild myalgia) were recorded at 20-21 days after immunization in a group 2 vaccinee, and fatigue was recorded at 28 days in a group 2 placebo recipient; after the second immunization in group 1, grade 2 dizziness and fatigue were recorded on day 7 in a vaccinee and grade 2 fatigue on day 7 in placebo recipient; after the third immunization in group 1, grade 2 headache was recorded at 14 days in a vaccinee. No systemic AEs were recorded in group 5. There were no severe/grade 3 solicited systemic AEs in any group. In summary, solicited AEs appeared balanced between vaccine and placebo groups and likely represented background rates in the community.

*Unsolicited AEs:* These were assessed by asking participant or parents "any other symptoms," described syndromically (e.g., "upper respiratory infection") and assessed for severity and additionally for relatedness to immunization (possibly, probably and definitely related combined as "related' and unlikely related and unrelated combined as "unrelated"). Unsolicited AEs occurred in 10/50 (20%) vaccinees and 3/25 (12%) placebo recipients (p=0.524) (**Table 1**), mild in severity except episodes of moderate abdominal pain, conjunctivitis and oropharyngeal pain in three adult vaccinees, moderate torticollis in one adult placebo recipient, and moderate furuncle in one group 3 vaccinee. All were deemed unrelated to investigational product and resolved without sequelae.

*Laboratory abnormalities:* Laboratory tests were white blood cell count, neutrophil count, lymphocyte count, platelet count, hemoglobin level, alanine aminotransferase (ALT), aspartate aminotransferase (AST) and creatinine. These were recorded 7 and 28 days after immunization, classified by pre-specified severity ranges and rated as clinically significant (CS) or not clinically significant (NCS). Overall, laboratory abnormalities occurred in 16/50 (32%) vaccinees and 14/25 (56%) placebo recipients (p=0.08) (**Table 1**). At day 7 post-vaccination, 7/50 (14%) vaccinees and 7/25 (28%) placebo recipients had abnormalities (p=0.142). These were grade 1 in severity except for grade 2 lymphocyte increases in two group 3 vaccinees and a grade 2 platelet decrease in a group 3 vaccinee. Additionally, one group 1 vaccinee had a grade 2 neutrophil decrease after the second vaccination. At day 28 post vaccination, 16/50 (32%) vaccinees and 11/25 (44%) placebo recipients had lab abnormalities (p=0.307), grade 1 except for one grade 2 neutrophil decrease in a group 1 vaccinee after the first immunization, two grade 2 lymphocyte increases and one grade 2 platelet decrease in group 3 vaccinees, one grade 2 lymphocyte increase in a group 4 placebo recipient, and one grade 2 platelet decrease in a group 5 placebo recipient. Another group 1 vaccinee had a grade 2 neutrophil decrease 28 days after the second vaccination. None of these abnormalities was deemed clinically significant. There was one grade 3 hemoglobin decrease in a group 4 vaccinee (#82) on day 28 after vaccination. This represented a 0.6 g/dL drop compared with the pre-immunization baseline of 9.6 g/dL and this shift was deemed not clinically significant. These various abnormalities appeared unrelated to vaccine administration with respect to timing, did not indicate differences between vaccinees and placebo recipients and likely represented background rates in the community.

<u>Long-term Safety Follow-up (at 12 and 24 weeks)</u>

Participants were scheduled for visits at 12 and 24 weeks after immunization (after third immunization in group 1) to review medical history and obtain blood samples for immunology; 72/75 (96%) participants were seen at 12 weeks and 70/75 (93%) at 24 weeks. In addition to these scheduled visits, participants were encouraged to contact the GRAS clinical team 24/7 for unscheduled visits in case of any medical concerns. At the scheduled 12 week visit, two participants (#81, #82) both administered vaccine in group 4 presented with clinical malaria, were diagnosed with Pf by thick blood smear, were treated with DHA-PP and had blood drawn for hematology testing, recording grade 3 hemoglobin levels, representing 0.7 and 0.9 g/dL drops from baseline, changes not deemed clinically significant. At the same visit, a participant receiving vaccine in group 3 (#101) presented with malaria symptoms and was similarly diagnosed and treated for Pf clinical malaria. At the scheduled 24 week visit, a placebo recipient in group 2 (#34) presented with Pf clinical malaria and dengue IgM positivity, had blood drawn for hematology testing and recorded a grade 3 platelet level of 88,000/µL deemed clinically significant that resolved following malaria treatment and repeat testing. Regarding unscheduled visits, a participant receiving vaccine (#131) presented 15 weeks after immunization with abdominal discomfort and had a decreased lymphocyte count (1600/µL) and mildly increased neutrophil count (8890/µL). This presentation was considered clinically significant and resolved after empiric treatment for intestinal helminth infection. A participant receiving vaccine (#101) presented with abdominal pain, jaundice and grade 3 elevations in ALT (209 IU/L) and AST (261 IU//L) 29 weeks after immunization with negative hepatitis serologies and was followed beyond the last scheduled study visit until complete resolution of the elevated liver function tests was documented. No particular patterns of illnesses related to investigational product could be discerned among these or other unscheduled visits.

<u>Malaria Infections After Completing 28 Days of Follow-up (details)</u>

*Cross sectional prevalence surveys:* The protocol was amended to allow cross-sectional prevalence surveys of the study groups at the 12- and 24- month follow-up visits. However, approval was not obtained in time to conduct several of these surveys. **Table S2** provides the survey results.

**Table S2: Twelve- and 24-week Prevalence Surveys**

| Group | Age (yrs) | # doses / #PfSPZ | # positive / # surveyed (percent) | | Parasites/µL |
|---|---|---|---|---|---|
| | | | 12 week survey | 24 week* survey | |
| 1 | 20-50 | 3 / $2x10^5$ | ND | NS: 1/5 (20%)<br>L2: 0/9 (0%) | 1480<br>- |
| 2 | 20-50 | 1 / $4x10^5$ | ND | ND | - |
| 3 | 11-19 | 1 / $2x10^5$ | ND | NS: 0/5 (0%)<br>L2: 3/8 (37.5%) | -<br>290, 1040, 9920 |
| 4 | 6-10 | 1 / $1x10^5$ | ND | NS: 1/5 (20%)<br>L2: 3/10 (30%) | 2642<br>47, 78, 193 |
| 5 | 1-5 | 1 / $1x10^5$ | NS: 0/5 (0%)<br>L2: 2/10 (20%) | NS: 1/4 (25%)<br>L2: 0/9 (0%) | 24633<br>283, 2800 |
| 3-5 | 1-19 | | NS: 0/5 (0%)<br>L2: 2/10 (20%) | NS: 2/14 (14.3%)<br>L2: 6/27 (22.2%) | |
| Overall | 1-50 | | NS: 0/5 (0%)<br>L2: 2/10 (20%) | NS: 3/19 (15.1%)<br>L2: 6/36 (16.7%) | |

NS = normal saline; L2 = PfSPZ-LARC2 Vaccine; ND = not done (protocol amendment not approved in time)

* 24 weeks post V3 in group 1

The numbers of positive TBS and overall sample size were too low to determine if vaccination induced protection. There were no discernable patterns suggesting that vaccination with a single dose offered protection. One survey was conducted in the three-dose group (24-week survey in group 1). In this survey, none of the vaccine recipients were positive, while 1/5 (20%) of normal saline recipients was positive.

*Other malaria infections:* At sporadic timepoints post-terminal treatment 28 days after the last vaccination, participants presented to the clinical team with symptoms suggestive of malaria. A TBS was made and read in real time. The list of participants found to be positive with symptomatic infections is provided in **Table S3**.

**Table S3: List of Clinical Malaria Cases Outside of the 28-day Immunization Period**

| Group | Age range (yr) | # Doses | ID | Treatment | Day positive* | Parasites /μL by TBS | Visit type |
|---|---|---|---|---|---|---|---|
| 1 | 20-50 | 3 | 008 | PfSPZ | 209 | 2,050 | unscheduled |
| | | | 017 | PfSPZ | 142 | 2,905 | unscheduled |
| 2 | 20-50 | 1 | 34 | NS | 165 | 1,242 | unscheduled |
| 3 | 11-19 | 1 | 107 | NS | 94 | 596,310 | unscheduled |
| | | | 101 | PfSPZ | 84 | 1,256 | unscheduled |
| 4 | 6-10 | 1 | 083 | NS | 75 | 272,225 | unscheduled |
| | | | 087 | NS | 159 | 430,230 | unscheduled |
| | | | 088 | PfSPZ | 81 | 99,613 | unscheduled |
| | | | 81 | PfSPZ | 83 | 14,051 | unscheduled |
| | | | 82 | PfSPZ | 84 | 2,557 | unscheduled |
| 5 | 1-5 | 1 | 141 | PfSPZ | 72 | 2,482 | unscheduled |
| Total infections in placebo recipients: 4<br>Total infections in vaccine recipients: 7 | | | | | | | |
| * days post V1 | | | | | | | |

Counting all malaria infections diagnosed post-terminal treatment 28 days after vaccination, there were 15 infections in 14 of 50 vaccinees (28%) and 7 infections in 7 of 25 placebo recipients (28%). Participant 101 (group 3 vaccine recipient, main subgroup) had two infections, the first resulting in an unscheduled visit 84 days after immunization (**Table S3**), and the second at the 24-week prevalence survey (**Table S2**). Other participants with malaria infection had only one.

Supplemental Immunogenicity Data

| Group | Age (years) | Treatment | N (max sero-converters) | Median (range) Net OD 1.0 | | | | | | | |
|---|---|---|---|---|---|---|---|---|---|---|---|
| | | | | Baseline OD 1.0 | V1 +14 days | V1 +28 days | V1 +84 days | V3 | V3 +14 days | V3 +28 days | V3 +84 days |
| 1 | 20-45 | $2x10^5$PfSPZ x3 | 10 (2) | 1,650 (657 - 2642) | NSC | | (same as V3+28) | 2,780 (2,780-2,780) | 4,873 (3,550-6,196) | 4,920 (3,994-5,846) | 4,830 (2,242-7,417) |
| | | NS x3 | 5 (0) | NSC | NSC | | (same as V3+28) | | | | |
| 2 | 20-45 | $4x10^5$PfSPZ x1 | 10 (3) | 1,099 (401 – 2,983) | 5,877 (5,877 – 5,877) | 3,889 (811 – 6,727) | NSC | | | | |
| | | NS x1 | 5 (0) | NSC | NSC | NSC | NSC | | | | |
| 3 | 11-19 | $2x10^5$PfSPZ x1 | 10 (4) | 472 (103 - 891) | 1,088 (347 – 6,564) | 5,738 (978 – 9,947) | 1,422 (1,422 – 1,422) | | | | |
| | | NS x1 | 5 (0) | NSC | NSC | NSC | NSC | | | | |
| 4 | 6-10 | $1x10^5$PfSPZ x1 | 10 (3) | 41 (1 - 159) | 316 (269 - 351) | 220 (130 – 6,087) | 398 (152 – 3,571) | | | | |
| | | NS x1 | 5 (1) | 123 (123 - 123) | NSC | NSC | 325 (325-325) | | | | |
| 5 | 1-5 | $1x10^5$PfSPZ x1 | 10 (6) | 44 (1 - 233) | 296 (71 – 1,568) | 193 (62 - 635) | 75 (70-80) | | | | |
| | | NS x1 | 5 (0) | NSC | NSC | NSC | NSC | | | | |

**Table S4**. **Net OD 1.0 in vaccinees who seroconverted.** Median (range) of net OD 1.0 IgG antibodies to PfCSP only in those who seroconverted. PfSPZ=PfSPZ-LARC2 Vaccine, NS=normal saline, NSC=no seroconverters, shaded cell = no blood collected or timepoint not relevant.

| Group | Age (years) | Treatment | N (max sero-converters) | Median (range) Net OD Ratio | | | | | | | |
|---|---|---|---|---|---|---|---|---|---|---|---|
| | | | | Baseline OD 1.0 | V1 +14 days | V1 +28 days | V1 +84 days | V3 | V3 +14 days | V3 +28 days | V3 +84 days |
| 1 | 20-45 | 2x10$^5$PfSPZ x3 | 10 (2) | 1,650 (657 – 2642) | NSC | | (same as V3+28) | 5.5 (5.2-5.5) | 4.85 (3.3-6.4) | 5.15 (3.2-7.1) | 4.1 (3.8-4.4) |
| | | NS x3 | 5 (0) | NSC | NSC | | (same as V3+28) | | | | |
| 2 | 20-45 | 4x10$^5$PfSPZ x1 | 10 (3) | 1,099 (401 – 2,983) | 6.3 (6.3-6.3) | 3.3 (3.0-4.5) | NSC | | | | |
| | | NS x1 | 5 (0) | NSC | NSC | NSC | NSC | | | | |
| 3 | 11-19 | 2x10$^5$PfSPZ x1 | 10 (4) | 472 (103 - 891) | 4.4 (3.3-8.4) | 5.4 (3.1-15.3) | 4.0 (4.0-4.0) | | | | |
| | | NS x1 | 5 (0) | NSC | NSC | NSC | NSC | | | | |
| 4 | 6-10 | 1x10$^5$PfSPZ x1 | 10 (3) | 41 (1 - 159) | 21.7 (6.2-317.0) | 90.5 (11.0-221.0) | 153.5 (3.5-275.7) | | | | |
| | | NS x1 | 5 (1) | 123 (123 - 123) | NSC | NSC | 3.6 (3.6 - 3.6) | | | | |
| 5 | 1-5 | 1x10$^5$PfSPZ x1 | 10 (6) | 44 (1 - 233) | 39.9 (4.0-265.0) | 117 (3.7-217.0) | 76 (71-81) | | | | |
| | | NS x1 | 5 (0) | NSC | NSC | NSC | NSC | | | | |

**Table S5**. **OD 1.0 ratio in vaccinees who seroconverted.** Median (range) of OD 1.0 ratios for IgG antibodies to PfCSP only in those who seroconverted. PfSPZ=PfSPZ-LARC2 Vaccine, NS=normal saline, NSC=no seroconverters, shaded cell = no blood collected or timepoint not relevant.

| Group | Age (years) | Treatment | N | Median (range) Net OD 1.0 | | | | | | | |
|---|---|---|---|---|---|---|---|---|---|---|---|
| | | | | Baseline | V1 +14 days | V1 +28 days | V1 +84 days | V3 | V3 +14 days | V3 +28 days | V3 +84 days |
| 1 | 20-45 | $2x10^5$PfSPZ x3 | 10 | 2,519 (290-11,219) | 36 (1-1,970) | | (same as V3+28) | 948 (1-3,932) | 1,395 (73-6,196) | 2,749 (357-12,142) | 1,857 (1-7,417) |
| | | NS x3 | 5 | 3,005 (481-4,273) | 1 (1-43) | | (same as V3+28) | 1 (1-1) | 1 (1-1) | 14 (1-1,131) | 1 (1-1) |
| 2 | 20-45 | $4x10^5$PfSPZ x1 | 10 | 1,344 (401-8,844) | 1,145 (1-5,877) | 1,775 (582-6,727) | 1,068 (226-2,491) | | | | |
| | | NS x1 | 5 | 2,270 (967-2,552) | 1 (1-231) | 1 (1-229) | 73 (1-446) | | | | |
| 3 | 11-19 | $2x10^5$PfSPZ x1 | 10 | 424 (103-891) | 66 (1-6,564) | 564 (1-9,947) | 1 (1-479) | | | | |
| | | NS x1 | 5 | 314 (175-563) | 23 (1-295) | 187 (1-416) | 20 (1-479) | | | | |
| 4 | 6-10 | $1x10^5$PfSPZ x1 | 10 | 168 (1-5,399) | 161 (1-1,831) | 140 (1-6,087) | 278 (1-3,571) | | | | |
| | | NS x1 | 5 | 91 (62-426) | 1 (1-17) | 22 (1-59) | 62 (1-635) | | | | |
| 5 | 1-5 | $1x10^5$PfSPZ x1 | 10 | 94 (1-909) | 175 (1-1,568) | 116 (1-635) | 1 (1-80) | | | | |
| | | NS x1 | 5 | 55 (45-16,045) | 53 (10-6,247) | 36 (1-5,825) | 1 (1-14) | | | | |

**Table S6. Net OD 1.0 in all vaccinees and controls**: Median (range) of net OD 1.0 IgG antibodies to PfCSP including all participants in the group. PfSPZ=PfSPZ-LARC2 Vaccine, NS=normal saline, shaded cell = no blood collected or timepoint not relevant.

| Group | Age (years) | Treatment | N | Median (range) Net OD 1.0 | | | | | | | |
|---|---|---|---|---|---|---|---|---|---|---|---|
| | | | | Baseline | V1 +14 days | V1 +28 days | V1 +84 days | V3 | V3 +14 days | V3 +28 days | V3 +84 days |
| 1 | 20-45 | $2x10^5$PfSPZ x3 | 10 | 2,519 (290-11,219) | 1.1 (1.0-1.7 | | (same as V3+28) | 1.6 (1.0-5.2) | 1.8 (1.0-6.4) | 2.1 (1.2-7.1) | 1.7 (1.0-4.4) |
| | | NS x3 | 5 | 3,005 (481-4,273) | 1.0 (1.0-1.0) | | (same as V3+28) | 1 .0 (1.0 – 1.0) | 1 .0 (1.0 – 1.0) | 1.0 (1.0 - 1.5) | 1.0 (1.0 – 1.0) |
| 2 | 20-45 | $4x10^5$PfSPZ x1 | 10 | 1,344 (401-8,844) | 1.8 (1.0 - 6.3) | 1.95 (1.2 - 4.5) | 1.7 (1.1 - 2.1) | | | | |
| | | NS x1 | 5 | 2,270 (967-2,552) | 1.0 (1.0 - 1.2) | 1.0 (1.0 - 1.2) | 1.0 (1.0 - 1.2) | | | | |
| 3 | 11-19 | $2x10^5$PfSPZ x1 | 10 | 424 (103-891) | 1.2 (1.0 - 8.4) | 2.55 (1.0 - 15.3) | 1.0 (1.0 – 4.0) | | | | |
| | | NS x1 | 5 | 314 (175-563) | 1.1 (1 - 1.5) | 1.5 (1.0 - 2.9) | 1.1 (1.0 - 1.9) | | | | |
| 4 | 6-10 | $1x10^5$PfSPZ x1 | 10 | 168 (1-5,399) | 1.6 (1.0 – 317.0) | 1.7 (1.0 – 221.0) | 2.3 (1.0 - 275.7) | | | | |
| | | NS x1 | 5 | 91 (62-426) | 1.0 (1.0 - 1.3) | 1.1 (1.0 - 1.7) | 1.7 (1.0 - 3.6) | | | | |
| 5 | 1-5 | $1x10^5$PfSPZ x1 | 10 | 94 (1-909) | 5.0 (1.0 – 265.0) | 2.55 (1.0 – 217.0) | 1 (1 - 81) | | | | |
| | | NS x1 | 5 | 55 (45-16,045) | 1.4 (1.2 - 2.2) | 1.4 (1 - 1.8) | 1 (1 – 1.3) | | | | |

**Table S7. OD 1.0 ratio in all vaccinees and controls**: Median (range) of OD 1.0 ratios for IgG antibodies to PfCSP including all participants in the group. PfSPZ=PfSPZ-LARC2 Vaccine, NS=normal saline, shaded cell = no blood collected or timepoint not relevant.

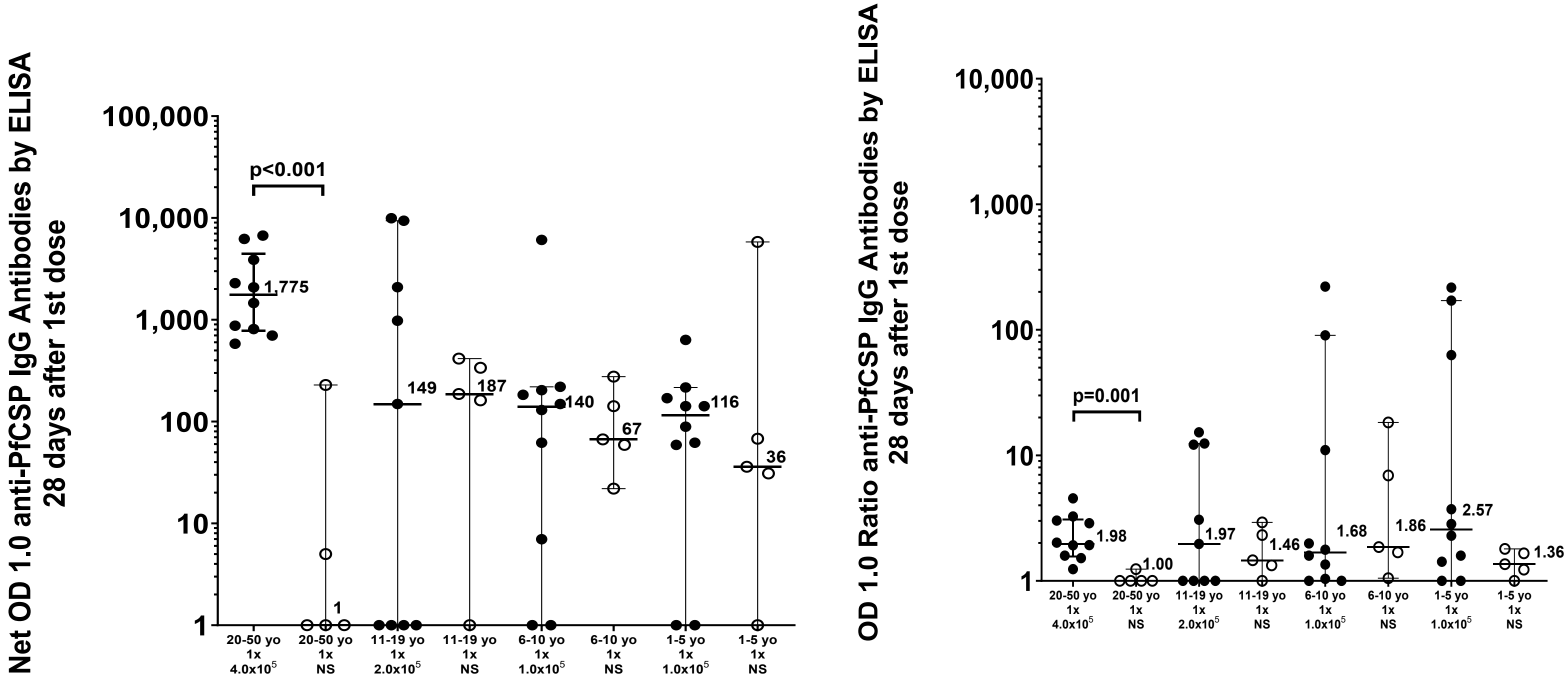


**Fig. S3. IgG Net Optical Density (OD 1.0) and IgG OD 1.0 ratio at 28 days in recipients who received a single dose:** Median and interquartile range of net optical density (OD) at 1.0 for IgG antibodies against PfCSP, measured by ELISA, 28 days after the first dose of either PfSPZ-LARC2 Vaccine or normal saline in BFSPZL1 participants. In both panels, filled black circles (●) represent vaccinees, and open black circles (○) indicate individuals who received Normal Saline.

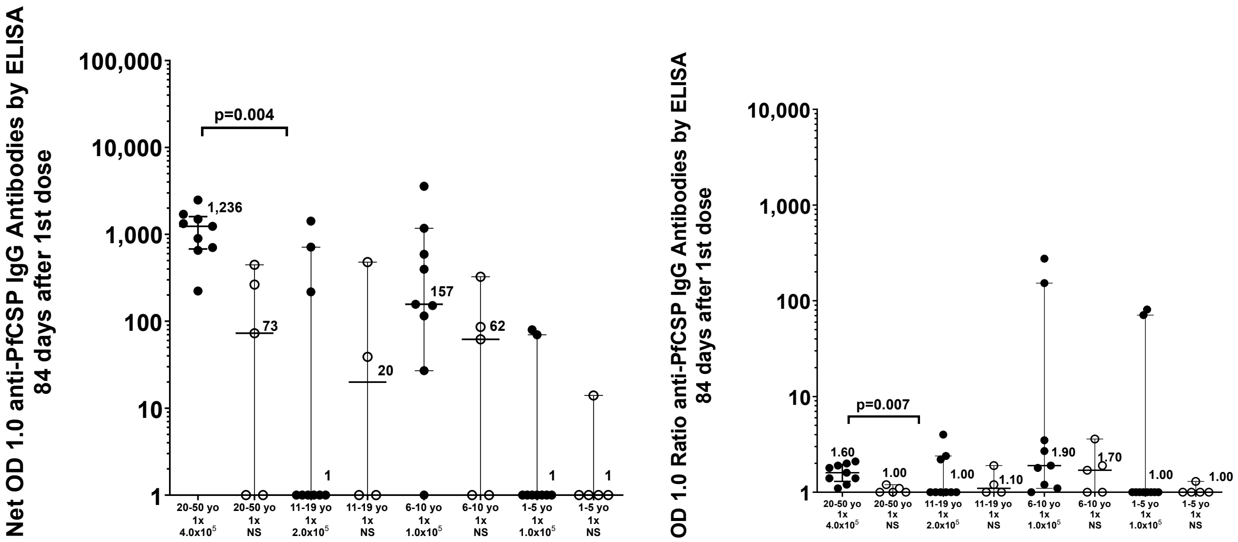


**Fig. S4. IgG Net Optical Density (OD 1.0) and IgG OD 1.0 ratio at 84 days in recipients who received a single dose:** Median and interquartile range of net optical density (OD) at 1.0 for IgG antibodies against PfCSP, measured by ELISA, 84 days after the first dose of either PfSPZ-LARC2 Vaccine or normal saline in BFSPZL1 participants. In both panels, filled black circles (●) represent vaccinees, and open black circles (○) indicate individuals who received Normal Saline.

Immunogenicity (detailed description):

IgG, IgM, and IgA antibodies to the Pf circumsporozoite protein (CSP) were assessed by enzyme-linked immunosorbent assay (ELISA) at baseline and at defined time points after immunization. The reciprocal serum dilution at which the optical density was 1.0 (OD 1.0) was calculated at each time point. Individuals were considered to have seroconverted if the post-immunization OD 1.0 minus the pre-immunization OD 1.0 (net OD 1.0) was ≥50 and the post-immunization OD 1.0 / pre-immunization OD 1.0 (OD 1.0 ratio) was ≥3.0. The antibody analysis included all 10 vaccinees and 5 controls in each group, although later timepoints were missed in dropouts.

*Baseline IgG antibodies to PfCSP:* Prior to immunization, many participants had high baseline levels of IgG antibodies to PfCSP as assessed by OD 1.0 (**Table 2**). The median baseline OD 1.0 was associated with age: 2,054 in 20-48-year-olds, 355 in 11-19 year olds, 151 in 6-10-year-olds, and 77 in 1-5-year-olds (Spearman's r=-0.739, p<0.0001). Despite the age-related trend, there was considerable variation in baseline OD 1.0 levels. For example, the highest baseline OD 1.0s in each group were 11,219 (20-48-year-olds), 891 (11-19-year-olds), 5,399 (6-10-year-olds), and 16,045 (1-5-year-olds).

*Individual seroconversion rates of IgG antibodies to PfCSP*: Seroconversion rates for IgG antibody to the PfCSP differed by age, dose, and time post immunization (**Table 2**). Seroconversion rates at 14 days after a single dose of PfSPZ-LARC2 were 0% in group 1 ($2.0\times10^5$ PfSPZ) and 10% in group 2 ($4.0\times10^5$ PfSPZ), 30% in group 3 ($2.0\times10^5$ PfSPZ), 30% in group 4 ($1.0\times10^5$ PfSPZ), and 60% in group 5 ($1.0\times10^5$ PfSPZ). No controls seroconverted at 14 days of immunization. Only the 60% seroconversion rate in 1-5-year-olds was statistically significant compared to the group's controls (p=0.042, Fisher's exact test, 2-tailed). Seroconversion rates at 28 days after a single dose were 30% in group 2, 40% in group 3, 30% in group 4, and 40% in group 5 (Table 2, Fig. S2). No controls seroconverted at 28 days after immunization. Seroconversion rates at 84 days after a single dose were 0% in group 2, 10% in group 3, 30% in group 4, and 20% in group 5. One control seroconverted at 84 days after immunization. Two of 10 (20%) vaccinees in group 1 seroconverted after the third dose of $2.0\times10^5$ PfSPZ. In these two individuals, anti-PfCSP antibodies were present at 14, 28, and 84 days after the 3rd dose of PfSPZ-LARC2 Vaccine. No vaccinees met criteria for seroconversion of IgM or IgA PfCSP antibodies (**Table S6)**.

*Magnitude and kinetics of anti-PfCSP antibodies in 3 dose group:* Group 1 IgG responses were significantly higher in vaccine recipients than in controls. At 14 days after the third dose, median net OD 1.0 was 1,395 vs 1 in controls (p=0.001, Mann-Whitney test) and median OD ratio was 1.8 vs 1.0 in controls (p = 0.004, Mann-Whitney test) (**Fig. 5**). The median net OD 1.0 rose from 36 at 14 days after the first dose to 1,395 at 14 days after the third dose (1,395 vs 36, p = 0.007) to 2,749 at 28 days after the third dose (2,749 vs 36, p = 0.001). Likewise, the median OD ratio rose from 1.05 at 14 days after the first dose to 1.8 at 14 days after the third dose (1.05 vs 1.8, p = 0.007) to 2.05 at 28 days after the third dose (2.05 vs 1.05, p < 0.001).

In group 2, IgG responses were also significantly higher in vaccine recipients than in controls. At 14 days after immunization, median net OD 1.0 was 1,145 vs 1.0 (p = 0.016) and the median OD ratio was 1.82 vs 1.0 (p = 0.016) (**Fig. 4**). At 28 days after the first dose median net OD 1.0 was 1,775 vs 1.0 (p < 0.001) and median OD ratio was 1.98 vs 1.0 (p = 0.001) (**Fig. S3**). At 84 days after the first dose median net OD 1.0 was 1,236 vs 73 (p = 0.004) and median OD ratio was 1.6 vs 1.0 (p = 0.007) (**Fig. S4**). In groups 3, 4 and 5, median responses were higher in vaccinees than in controls, but the differences were mostly not statistically significant. Median net OD 1.0s at 14 days were 66, 161 and 175 and at 28 days were 564, 140 and 116 in the three groups of vaccinees, respectively. Median OD 1.0 ratios at day 14 were 1.2, 1.6 and 5.0, and at day 28 were 2.55, 1.7 and 2.55, respectively. Among comparisons between vaccinees and controls in these three younger age groups, only OD 1.0 ratios in group 5 were significantly different from those in controls (p=0.04).

*Dose effects:* A single high dose of PfSPZ administered to adults in group 2 of $4.0\times10^5$ PfSPZ, elicited greater antibody responses at 14 days than the response in adults in group 1 who received $2.0\times10^5$ PfSPZ. The median net OD 1.0 for the adult recipients of $4.0\times10^5$ PfSPZ was 1,145 (range 1-5,877) and the median net OD 1.0 for the adult recipients of $2.0\times10^5$ PfSPZ was 36 (range 1-1,970) (p=0.047). The median OD 1.0 ratio for the adult recipients of $4.0\times10^5$ PfSPZ was 1.8 (range 1.0-6.3) and the median OD 1.0 ratio for the adult recipients of $2.0\times10^5$ PfSPZ was 1.1 (range 1.0-1.7) (p=0.009) (**Fig. 4**).

*Age-dependent anti-PfCSP antibodies:* An age-dependent pattern was observed. Despite the fact that adolescents in group 3 received $2.0\times10^5$ PfSPZ and children in groups 4 and 5 received $1.0\times10^5$ PfSPZ, the children had higher immune responses. At 14 days post-immunization, groups 4 and 5 had median net OD 1.0s nearly three times higher (161 and 175, respectively) than those in the adolescent group 3 (66), though differences were not statistically significant. OD 1.0 ratios 14 days post immunization were similarly higher with decreasing age at 1.2, 1.6, and 5.0 in 11-19-year-olds, 6-10-year-olds and 1-5-year-olds, respectively (**Fig. 4**). The 5.0 ratio in 1-5-year-olds was significantly higher than the 1.2 ratio in 11-19-year-olds (p=0.045, Mann Whitney) (**Fig. 4**).

*Comparison with Mali PfSPZ-CVac trial:* The 20% seroconversion rate following 3 doses of PfSPZ-LARC2 Vaccine administered at $2x10^5$ PfSPZ per dose every four weeks to malaria-exposed adults in group 1 was modest but comparable to the 28% rate reported in malaria-exposed Malian adults after 3 doses of PfSPZ-CVac (chloroquine), administered every four weeks at a similar dose ($2.048 \times 10^5$ PfSPZ) (Coulibaly, eClinicalMedicine 2022). Both PfSPZ-CVac (chloroquine) and PfSPZ-LARC2 Vaccine parasites replicate and should provide similar immunogenicity with respect to PfCSP. The low seroconversion rate observed in both trials likely reflects the generally attenuated immune responses to PfCSP in heavily malaria-exposed African populations. Interestingly, group 1 vaccine recipients exhibited an approximately 8-fold higher baseline OD 1.0 value for PfCSP antibodies (median of 2,519 and 3,005) (**Table 2**) compared with vaccine recipients in the Mali trial (median of 352) using the identical ELISA. To facilitate comparison between the two trials, we co-plotted median absolute OD 1.0 values. These plots show similar separations between vaccine and control groups, corresponding to similar (low) seroconversion rates, but overall higher values in BFSPZL1, reflecting the higher baseline antibody levels (**Fig. S5**). Net OD 1.0 values were also higher in BFSPZL1. For example, at the first post-V3 time point in the Mali trial (V3+12), the net median OD 1.0 was 414, calculated as 822 minus 408, (**Fig. S5**) compared with 2,018 at the corresponding post-V3 time point in BFSPZL1 (V3+14), calculated as 4,537 minus 2519 (**Fig. S5**). (Note that 2,018, a difference in the median values of two time points, is close to but different from the true net OD 1.0 at timepoint V3+14, which is 1,395 as presented in **Fig. 5**, calculated as the median of the individual net OD 1.0 values for each participant). OD 1.0 ratios were similar between the two trials: at V3+12 in Mali, OD 1.0 ratios were 1.63 (participants later infected with Pf) and 1.70 (participants not infected), and at V3+14 in Burkina Faso, the OD 1.0 ratio was 1.8.

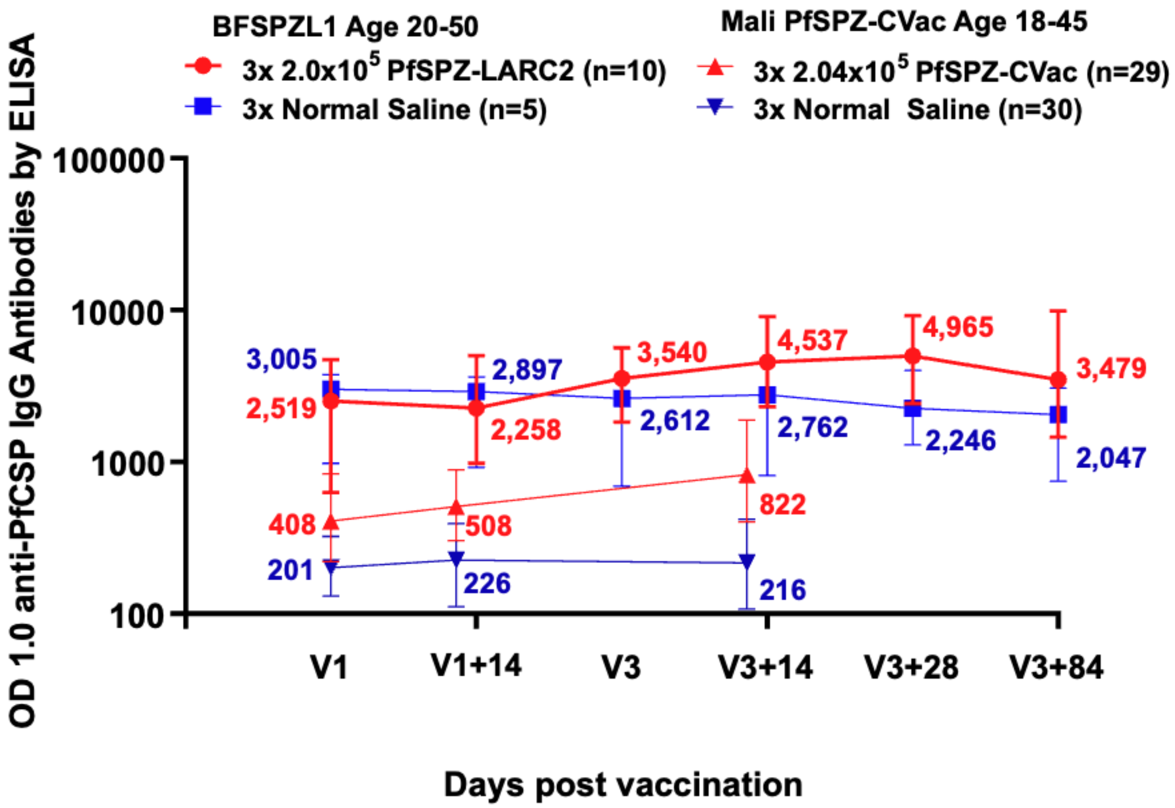


**Fig. S6: Anti-PfCSP Antibody Levels by ELISA:** Kinetics of median absolute OD 1.0 for IgG antibodies to PfCSP, shown with interquartile ranges by treatment group. The red lines represent anti-PfCSP ELISA OD 1.0 values for the vaccine groups in the BFSPZL1 and Mali PfSPZ-CVac trials, while the blue lines represent anti-PfCSP ELISA OD 1.0 values for the control groups.

**Table S5. All PfCSP IgG responses measured by ELISA in volunteers enrolled in the BFSPZL1 trial**: Antibody responses are reported as optical density (OD) 1.0 values for each volunteer by study group, age range, dose, treatment arm, and visit time point. Values below the assay range and zero values were assigned a value of 1 for tabulation and summary statistics. Net values represent the change from baseline at the indicated visit; ratios represent the fold change relative to baseline. NS indicates not sampled or not available. Median, mean, and geometric mean rows summarize responses within the corresponding treatment group.

A. <u>Group 1</u>

| Group | Age | Dose | ID | Treatment Arm Allocation | IgG ELISA PfCSP OD 1.0 | | | | | | | | | | | | | | | |
|---|---|---|---|---|---|---|---|---|---|---|---|---|---|---|---|---|---|---|---|---|
| | | | | | IgG ELISA PfCSP OD 1.0 | IgG ELISA PfCSP OD 1.0 V1+14 | | | IgG ELISA PfCSP OD 1.0 V3 | | | IgG ELISA PfCSP OD 1.0 V3+14 | | | IgG ELISA PfCSP OD 1.0 V3+28 | | | IgG ELISA PfCSP OD 1.0 V3+84 | | |
| | | | | | V1 | V1+14 | Net | Ratio | V3 | Net | Ratio | V3+14 | Net | Ratio | V3+28 | Net | Ratio | V3+84 | Net | Ratio |
| 1 | 20 - 50 | 3x 2.0x$10^5$ | BFSPZL-008 | PfSPZ-LARC2 Vaccine | 11,219 | 9,755 | 1 | 1 | 12,608 | 1,389 | 1 | 11,292 | 73 | 1 | 23,361 | 12,142 | 2 | 15,183 | 3,964 | 1 |
| 1 | 20 - 50 | 3x 2.0x$10^5$ | BFSPZL-013 | PfSPZ-LARC2 Vaccine | 657 | 1,102 | 445 | 2 | 3,437 | 2,780 | 5 | 4,207 | 3,550 | 6 | 4,651 | 3,994 | 7 | 2,899 | 2,242 | 4 |
| 1 | 20 - 50 | 3x 2.0x$10^5$ | BFSPZL-015 | PfSPZ-LARC2 Vaccine | 290 | 286 | 1 | 1 | 676 | 386 | 2 | 724 | 434 | 3 | 647 | 357 | 2 | 385 | 95 | 1 |
| 1 | 20 - 50 | 3x 2.0x$10^5$ | BFSPZL-017 | PfSPZ-LARC2 Vaccine | 535 | 606 | 71 | 1 | 1,301 | 766 | 2 | 1,296 | 761 | 2 | 1,277 | 742 | 2 | 927 | 392 | 2 |
| 1 | 20 - 50 | 3x 2.0x$10^5$ | BFSPZL-025 | PfSPZ-LARC2 Vaccine | 1,622 | 1,913 | 291 | 1 | 3,321 | 1,699 | 2 | 3,006 | 1,384 | 2 | 3,124 | 1,502 | 2 | 3,479 | 1,857 | 2 |
| 1 | 20 - 50 | 3x 2.0x$10^5$ | BFSPZL-036 | PfSPZ-LARC2 Vaccine | 3,462 | 2,602 | 1 | 1 | 4,356 | 894 | 1 | 4,867 | 1,405 | 1 | 5,278 | 1,816 | 2 | NS | NS | NS |
| 1 | 20 - 50 | 3x 2.0x$10^5$ | BFSPZL-037 | PfSPZ-LARC2 Vaccine | 4,418 | 4,171 | 1 | 1 | 4,122 | 1 | 1 | 6,049 | 1,631 | 1 | 8,099 | 3,681 | 2 | 4,493 | 75 | 1 |
| 1 | 20 - 50 | 3x 2.0x$10^5$ | BFSPZL-039 | PfSPZ-LARC2 Vaccine | 5,532 | 7,502 | 1,970 | 1 | 9,464 | 3,932 | 2 | 9,673 | 4,141 | 2 | 11,265 | 5,733 | 2 | 9,693 | 4,161 | 2 |
| 1 | 20 - 50 | 3x 2.0x$10^5$ | BFSPZL-056 | PfSPZ-LARC2 Vaccine | 2,642 | 3,272 | 630 | 1 | 3,643 | 1,001 | 1 | 8,838 | 6,196 | 3 | 8,488 | 5,846 | 3 | 10,059 | 7,417 | 4 |
| 1 | 20 - 50 | 3x 2.0x$10^5$ | BFSPZL-067 | PfSPZ-LARC2 Vaccine | 2,396 | 1,172 | 1 | 1 | 2,003 | 1 | 1 | 2,640 | 244 | 1 | 2,800 | 404 | 1 | 1,984 | 1 | 1 |
| **Median** | | | | | **2,519** | **2,258** | **36** | **1** | **3,540** | **948** | **2** | **4,537** | **1,395** | **2** | **4,965** | **2,749** | **2** | **3,479** | **1,857** | **2** |
| **Mean** | | | | | **3,277** | **3,238** | **341** | **1** | **4,493** | **1,285** | **2** | **5,259** | **1,982** | **2** | **6,899** | **3,622** | **3** | **5,456** | **2,245** | **2** |
| **GeoMean** | | | | | **1,994** | **2,017** | **20** | **1** | **3,308** | **304** | **2** | **3,935** | **1,054** | **2** | **4,473** | **2,110** | **2** | **3,309** | **483** | **2** |
| 1 | 20 - 50 | 3x NS | BFSPZL-014 | Normal Saline placebo | 1,472 | 1,515 | 43 | 1 | 999 | 1 | 1 | 1,268 | 1 | 1 | 2,246 | 774 | 2 | 1,038 | 1 | 1 |
| 1 | 20 - 50 | 3x NS | BFSPZL-016 | Normal Saline placebo | 481 | 315 | 1 | 1 | 383 | 1 | 1 | 359 | 1 | 1 | 495 | 14 | 1 | 450 | 1 | 1 |
| 1 | 20- 50 | 3x NS | BFSPZL-023 | Normal Saline placebo | 4,273 | 4,316 | 43 | 1 | 2,806 | 1 | 1 | 3,206 | 1 | 1 | 3,669 | 1 | 1 | 3,606 | 1 | 1 |
| 1 | 20 - 50 | 3x NS | BFSPZL-027 | Normal Saline placebo | 3,005 | 2,907 | 1 | 1 | 2,612 | 1 | 1 | 2,878 | 1 | 1 | 2,089 | 1 | 1 | 2,047 | 1 | 1 |
| 1 | 20 - 50 | 3x NS | BFSPZL-032 | Normal Saline placebo | 3,206 | 2,897 | 1 | 1 | 2,910 | 1 | 1 | 2,762 | 1 | 1 | 4,337 | 1,131 | 1 | 2,504 | 1 | 1 |
| **Median** | | | | | **3,005** | **2,897** | **1** | **1** | **2,612** | **1** | **1** | **2,762** | **1** | **1** | **2,246** | **14** | **1** | **2,047** | **1** | **1** |
| **Mean** | | | | | **2,487** | **2,390** | **18** | **1** | **1,942** | **1** | **1** | **2,095** | **1** | **1** | **2,567** | **384** | **1** | **1,929** | **1** | **1** |
| **GeoMean** | | | | | **1,963** | **1,769** | **5** | **1** | **1,522** | **1** | **1** | **1,633** | **1** | **1** | **2,058** | **26** | **1** | **1,539** | **1** | **1** |

B. <u>Groups 2-5</u>

| Group | Age | Dose | ID | Treatment Arm Allocation | IgG ELISA PfCSP OD 1.0 | | | | | | | | | |
|---|---|---|---|---|---|---|---|---|---|---|---|---|---|---|
| | | | | | IgG ELISA PfCSP OD 1.0 | IgG ELISA PfCSP OD 1.0 V1+14 | | | IgG ELISA PfCSP OD 1.0 V1+28 | | | IgG ELISA PfCSP OD 1.0 V1+84 | | |
| | | | | | **V1** | **V1+14** | **Net** | **Ratio** | **V1+28** | **Net** | **Ratio** | **V1+84** | **Net** | **Ratio** |
| 2 | 20 - 50 | 1x 4.0x$10^5$ | BFSPZL-011 | PfSPZ-LARC2 Vaccine | 8,844 | 7,159 | 1 | 1 | 10,930 | 2,086 | 1 | 10,080 | 1,236 | 1 |
| 2 | 20 - 50 | 1x 4.0x$10^5$ | BFSPZL-012 | PfSPZ-LARC2 Vaccine | 1,471 | 2,631 | 1,160 | 2 | 2,344 | 873 | 2 | 2,964 | 1,493 | 2 |
| 2 | 20 - 50 | 1x 4.0x$10^5$ | BFSPZL-029 | PfSPZ-LARC2 Vaccine | 6,116 | 6,112 | 1 | 1 | 12,348 | 6,232 | 2 | 8,607 | 2,491 | 1 |
| 2 | 20 - 50 | 1x 4.0x$10^5$ | BFSPZL-049 | PfSPZ-LARC2 Vaccine | 1,099 | 6,976 | 5,877 | 6 | 4,988 | 3,889 | 5 | 1,805 | 706 | 2 |
| 2 | 20 - 50 | 1x 4.0x$10^5$ | BFSPZL-058 | PfSPZ-LARC2 Vaccine | 1,216 | 2,540 | 1,324 | 2 | 3,507 | 2,291 | 3 | 1,439 | 223 | 1 |
| 2 | 20 - 50 | 1x 4.0x$10^5$ | BFSPZL-062 | PfSPZ-LARC2 Vaccine | 1,579 | 2,899 | 1,320 | 2 | 3,042 | 1,463 | 2 | 3,284 | 1,705 | 2 |
| 2 | 20 - 50 | 1x 4.0x$10^5$ | BFSPZL-063 | PfSPZ-LARC2 Vaccine | 759 | 1,889 | 1,130 | 3 | 1,458 | 699 | 2 | 1,413 | 654 | 2 |
| 2 | 20 - 50 | 1x 4.0x$10^5$ | BFSPZL-066 | PfSPZ-LARC2 Vaccine | 1,109 | 1,566 | 457 | 1 | 1,691 | 582 | 2 | 2,008 | 899 | 2 |
| 2 | 20 - 50 | 1x 4.0x$10^5$ | BFSPZL-069 | PfSPZ-LARC2 Vaccine | 401 | 871 | 470 | 2 | 1,212 | 811 | 3 | NS | NS | NS |
| 2 | 20 - 50 | 1x 4.0x$10^5$ | BFSPZL-070 | PfSPZ-LARC2 Vaccine | 2,983 | 4,721 | 1,738 | 2 | 9,710 | 6,727 | 3 | 4,312 | 1,329 | 1 |
| **Median** | | | | | **1,344** | **2,765** | **1,145** | **2** | **3,275** | **1,775** | **2** | **2,964** | **1,236** | **2** |
| **Mean** | | | | | **2,558** | **3,736** | **1,348** | **2** | **5,123** | **2,565** | **2** | **3,990** | **1,193** | **2** |
| **GeoMean** | | | | | **1,670** | **3,060** | **295** | **2** | **3,706** | **1,785** | **2** | **3,105** | **996** | **2** |
| 2 | 20 - 50 | 1x NS | BFSPZL-024 | Normal Saline placebo | 2,270 | 1,987 | 1 | 1 | 2,275 | 5 | 1 | 2,534 | 264 | 1 |
| 2 | 20 - 50 | 1x NS | BFSPZL-034 | Normal Saline placebo | 2,552 | 2,401 | 1 | 1 | 2,488 | 1 | 1 | 2,998 | 446 | 1 |
| 2 | 20 - 50 | 1x NS | BFSPZL-052 | Normal Saline placebo | 967 | 1,198 | 231 | 1 | 1,196 | 229 | 1 | 727 | 1 | 1 |
| 2 | 20 - 50 | 1x NS | BFSPZL-061 | Normal Saline placebo | 1,838 | 1,833 | 1 | 1 | 1,644 | 1 | 1 | 1,009 | 1 | 1 |
| 2 | 20 - 50 | 1x NS | BFSPZL-068 | Normal Saline placebo | 2,359 | 1,982 | 1 | 1 | 2,132 | 1 | 1 | 2,432 | 73 | 1 |
| **Median** | | | | | **2,270** | **1,982** | **1** | **1** | **2,132** | **1** | **1** | **2,432** | **73** | **1** |
| **Mean** | | | | | **1,997** | **1,880** | **47** | **1** | **1,947** | **47** | **1** | **1,940** | **157** | **1** |
| **GeoMean** | | | | | **1,893** | **1,834** | **3** | **1** | **1,884** | **4** | **1** | **1,684** | **24** | **1** |
| 3 | 11 – 19 | 1x 2.0x$10^5$ | BFSPZL-095 | PfSPZ-LARC2 Vaccine | 183 | 183 | 1 | 1 | **2,277** | 2,094 | 12 | 401 | 218 | 2 |
| 3 | 11 – 19 | 1x 2.0x$10^5$ | BFSPZL-099 | PfSPZ-LARC2 Vaccine | 103 | 450 | 347 | 4 | NS | NS | NS | NS | NS | NS |
| 3 | 11 – 19 | 1x 2.0x$10^5$ | BFSPZL-101 | PfSPZ-LARC2 Vaccine | 472 | 1,560 | 1,088 | 3 | 1,450 | 978 | 3 | 1,894 | 1,422 | 4 |
| 3 | 11 – 19 | 1x 2.0x$10^5$ | BFSPZL-105 | PfSPZ-LARC2 Vaccine | 657 | 690 | 33 | 1 | **10,039** | 9,382 | 15 | 272 | 1 | 1 |
| 3 | 11 – 19 | 1x 2.0x$10^5$ | BFSPZL-108 | PfSPZ-LARC2 Vaccine | 154 | 165 | 11 | 1 | 303 | 149 | 2 | 71 | 1 | 1 |
| 3 | 11 – 19 | 1x 2.0x$10^5$ | BFSPZL-110 | PfSPZ-LARC2 Vaccine | 891 | 7,455 | 6,564 | 8 | **10,838** | 9,947 | 12 | 184 | 1 | 1 |
| 3 | 11 – 19 | 1x 2.0x$10^5$ | BFSPZL-114 | PfSPZ-LARC2 Vaccine | 514 | 537 | 23 | 1 | 304 | 1 | 1 | 1,227 | 713 | 2 |
| 3 | 11 – 19 | 1x 2.0x$10^5$ | BFSPZL-115 | PfSPZ-LARC2 Vaccine | 375 | 423 | 48 | 1 | 162 | 1 | 1 | 311 | 1 | 1 |
| 3 | 11 – 19 | 1x 2.0x$10^5$ | BFSPZL-118 | PfSPZ-LARC2 Vaccine | 309 | 392 | 83 | 1 | 260 | 1 | 1 | 90 | 1 | 1 |
| 3 | 11 – 19 | 1x 2.0x$10^5$ | BFSPZL-121 | PfSPZ-LARC2 Vaccine | 855 | 1,587 | 732 | 2 | 29 | 1 | 1 | 805 | 1 | 1 |
| **Median** | | | | | **424** | **494** | **66** | **1** | **304** | **149** | **2** | **311** | **1** | **1** |
| **Mean** | | | | | **451** | **1,344** | **893** | **2** | **2,851** | **2,506** | **5** | **584** | **262** | **2** |
| **GeoMean** | | | | | **364** | **659** | **95** | **2** | **700** | **67** | **3** | **348** | **8** | **1** |
| 3 | 11 – 19 | 1x NS | BFSPZL-089 | Normal Saline placebo | 231 | 221 | 1 | 1 | **157** | 1 | 1 | 270 | 39 | 1 |
| 3 | 11 – 19 | 1x NS | BFSPZL-103 | Normal Saline placebo | 563 | 858 | 295 | 2 | 750 | 187 | 1 | 1,042 | 479 | 2 |
| 3 | 11 – 19 | 1x NS | BFSPZL-107 | Normal Saline placebo | 314 | 337 | 23 | 1 | 730 | 416 | 2 | 45 | 1 | 1 |
| 3 | 11 - 19 | 1x NS | BFSPZL-112 | Normal Saline placebo | 175 | 156 | 1 | 1 | 513 | 338 | 3 | NS | NS | NS |
| 3 | 11 - 19 | 1x NS | BFSPZL-116 | Normal Saline placebo | 355 | 512 | 157 | 1 | 517 | 162 | 2 | 173 | 1 | 1 |
| **Median** | | | | | **314** | **337** | **23** | **1** | **517** | **187** | **2** | **222** | **20** | **1** |
| **Mean** | | | | | **328** | **417** | **95** | **1** | **533** | **221** | **2** | **383** | **130** | **1** |
| **GeoMean** | | | | | **303** | **348** | **16** | **1** | **469** | **84** | **2** | **216** | **12** | **1** |

| | | | | | | | | | | | | | | |
|---|---|---|---|---|---|---|---|---|---|---|---|---|---|---|
| 4 | 6 - 10 | 1x 2.0x$10^5$ | BFSPZL-073 | PfSPZ-LARC2 Vaccine | 68 | 419 | 351 | 6 | 6,155 | 6,087 | 91 | NS | NS | NS |
| 4 | 6 - 10 | 1x 2.0x$10^5$ | BFSPZL-075 | PfSPZ-LARC2 Vaccine | 239 | 359 | 120 | 2 | 422 | 183 | 2 | 266 | 27 | 1 |
| 4 | 6 - 10 | 1x 2.0x$10^5$ | BFSPZL-081 | PfSPZ-LARC2 Vaccine | 159 | 234 | 75 | 2 | 166 | 7 | 1 | 557 | 398 | 4 |
| 4 | 6 - 10 | 1x 2.0x$10^5$ | BFSPZL-082 | PfSPZ-LARC2 Vaccine | 13 | 282 | 269 | 22 | 143 | 130 | 11 | 3,584 | 3,571 | 276 |
| 4 | 6 - 10 | 1x 2.0x$10^5$ | BFSPZL-084 | PfSPZ-LARC2 Vaccine | 5,399 | 7,230 | 1,831 | 1 | 3,636 | 1 | 1 | 6,575 | 1,176 | 1 |
| 4 | 6 - 10 | 1x 2.0x$10^5$ | BFSPZL-085 | PfSPZ-LARC2 Vaccine | 1 | 317 | 316 | 317 | 221 | 220 | 221 | 153 | 152 | 153 |
| 4 | 6 - 10 | 1x 2.0x$10^5$ | BFSPZL-088 | PfSPZ-LARC2 Vaccine | 343 | 407 | 64 | 1 | 547 | 204 | 2 | 933 | 590 | 3 |
| 4 | 6 - 10 | 1x 2.0x$10^5$ | BFSPZL-090 | PfSPZ-LARC2 Vaccine | 621 | 569 | 1 | 1 | 412 | 1 | 1 | 249 | 1 | 1 |
| 4 | 6 - 10 | 1x 2.0x$10^5$ | BFSPZL-119 | PfSPZ-LARC2 Vaccine | 176 | 299 | 123 | 2 | 238 | 62 | 1 | 333 | 157 | 2 |
| 4 | 6 - 10 | 1x 2.0x$10^5$ | BFSPZL-120 | PfSPZ-LARC2 Vaccine | 151 | 350 | 199 | 2 | 300 | 149 | 2 | 266 | 115 | 2 |
| | | | **Median** | | **168** | **355** | **161** | **2** | **356** | **140** | **2** | **333** | **157** | **2** |
| | | | **Mean** | | **717** | **1,047** | **335** | **36** | **1,224** | **704** | **33** | **1,435** | **687** | **49** |
| | | | **GeoMean** | | **126** | **472** | **123** | **4** | **490** | **58** | **4** | **606** | **161** | **5** |
| 4 | 6 - 10 | 1x NS | BFSPZL-074 | Normal Saline placebo | 426 | 338 | 1 | 1 | 448 | 22 | 1 | 408 | 1 | 1 |
| 4 | 6 - 10 | 1x NS | BFSPZL-079 | Normal Saline placebo | 62 | 79 | 17 | 1 | 91 | 29 | 2 | 1 | 1 | 1 |
| 4 | 6 - 10 | 1x NS | BFSPZL-080 | Normal Saline placebo | 123 | 92 | 1 | 1 | 69 | 1 | 1 | 448 | 325 | 4 |
| 4 | 6 - 10 | 1x NS | BFSPZL-083 | Normal Saline placebo | 91 | 25 | 1 | 1 | 1 | 1 | 1 | 177 | 86 | 2 |
| 4 | 6 - 10 | 1x NS | BFSPZL-087 | Normal Saline placebo | 86 | 54 | 1 | 1 | 145 | 59 | 2 | 148 | 62 | 2 |
| | | | **Median** | | **91** | **79** | **1** | **1** | **91** | **22** | **1** | **177** | **62** | **2** |
| | | | **Mean** | | **158** | **118** | **4** | **1** | **151** | **22** | **1** | **236** | **95** | **2** |
| | | | **GeoMean** | | **121** | **80** | **2** | **1** | **53** | **8** | **1** | **86** | **18** | **2** |
| 5 | 1 - 5 | 1x 2.0x$10^5$ | BFSPZL-122 | PfSPZ-LARC2 Vaccine | 909 | 687 | 1 | 1 | 483 | 1 | 1 | 217 | 1 | 1.00 |
| 5 | 1 - 5 | 1x 2.0x$10^5$ | BFSPZL-130 | PfSPZ-LARC2 Vaccine | 1 | 265 | 264 | 265 | 171 | 170 | 171 | 71 | 70 | 71.00 |
| 5 | 1 - 5 | 1x 2.0x$10^5$ | BFSPZL-131 | PfSPZ-LARC2 Vaccine | 1 | 72 | 71 | 72 | 217 | 216 | 217 | 1 | 1 | 1.00 |
| 5 | 1 - 5 | 1x 2.0x$10^5$ | BFSPZL-132 | PfSPZ-LARC2 Vaccine | 77 | 463 | 386 | 6 | 219 | 142 | 3 | 70 | 1 | 1.00 |
| 5 | 1 - 5 | 1x 2.0x$10^5$ | BFSPZL-134 | PfSPZ-LARC2 Vaccine | 110 | 437 | 327 | 4 | 252 | 142 | 2 | 78 | 1 | 1.00 |
| 5 | 1 - 5 | 1x 2.0x$10^5$ | BFSPZL-136 | PfSPZ-LARC2 Vaccine | 233 | 1,801 | 1,568 | 8 | 868 | 635 | 4 | 200 | 1 | 1.00 |
| 5 | 1 - 5 | 1x 2.0x$10^5$ | BFSPZL-141 | PfSPZ-LARC2 Vaccine | 1 | 109 | 108 | 109 | 63 | 62 | 63 | 81 | 80 | 81.00 |
| 5 | 1 - 5 | 1x 2.0x$10^5$ | BFSPZL-144 | PfSPZ-LARC2 Vaccine | 76 | 107 | 31 | 1 | 1 | 1 | 1 | 13 | 1 | 1.00 |
| 5 | 1 - 5 | 1x 2.0x$10^5$ | BFSPZL-146 | PfSPZ-LARC2 Vaccine | 140 | 271 | 131 | 2 | 199 | 59 | 1 | 50 | 1 | 1.00 |
| 5 | 1 - 5 | 1x 2.0x$10^5$ | BFSPZL-147 | PfSPZ-LARC2 Vaccine | 152 | 371 | 219 | 2 | 241 | 89 | 2 | 45 | 1 | 1.00 |
| | | | **Median** | | **94** | **321** | **175** | **5** | **218** | **116** | **3** | **71** | **1** | **1.00** |
| | | | **Mean** | | **170** | **458** | **311** | **47** | **271** | **152** | **46** | **83** | **16** | **15.80** |
| | | | **GeoMean** | | **35** | **298** | **114** | **9** | **138** | **52** | **6** | **46** | **2** | **2.00** |
| 5 | 1 - 5 | 1x NS | BFSPZL-124 | Normal Saline placebo | 16,045 | 22,292 | 6,247 | 1 | 21,870 | 5,825 | 1 | 10241 | 1 | 1.00 |
| 5 | 1 - 5 | 1x NS | BFSPZL-135 | Normal Saline placebo | 298 | 404 | 106 | 1 | 366 | 68 | 1 | 192 | 1 | 1.00 |
| 5 | 1 - 5 | 1x NS | BFSPZL-140 | Normal Saline placebo | 47 | 88 | 41 | 2 | 78 | 31 | 2 | 1 | 1 | 1.00 |
| 5 | 1 - 5 | 1x NS | BFSPZL-143 | Normal Saline placebo | 55 | 65 | 10 | 1 | 53 | 1 | 1 | 69 | 14 | 1.30 |
| 5 | 1 - 5 | 1x NS | BFSPZL-149 | Normal Saline placebo | 45 | 98 | 53 | 2 | 81 | 36 | 2 | 17 | 1 | 1.00 |
| | | | **Median** | | **55** | **98** | **53** | **1** | **81** | **36** | **1** | **69** | **1** | **1.00** |
| | | | **Mean** | | **3,298** | **4,589** | **1,291** | **2** | **4,490** | **1,192** | **1** | **2104** | **4** | **1.06** |
| | | | **GeoMean** | | **223** | **347** | **108** | **2** | **306** | **54** | **1** | **75** | **2** | **1.05** |

**Table S6. All PfCSP IgM and IgA responses measured by ELISA in volunteers enrolled in the BFSPZL1 trial**: Antibody responses are reported as optical density (OD) 1.0 values for each volunteer by study group, age range, dose, treatment arm, and visit time point. Values below the assay range and zero values were assigned a value of 1 for tabulation and summary statistics. Net values represent the change from baseline at the indicated visit; ratios represent the fold change relative to baseline. NS indicates not sampled or not available. Median, mean, and geometric mean rows summarize responses within the corresponding treatment group.

A. <u>Group 1 – PfCSP IgM</u>

| Group | Age | Dose | ID | Treatment Arm Allocation | IgM ELISA PfCSP OD 1.0 | | | | | | | | | | | | | | | |
|---|---|---|---|---|---|---|---|---|---|---|---|---|---|---|---|---|---|---|---|---|
| | | | | | IgM ELISA PfCSP OD 1.0 | IgM ELISA PfCSP OD 1.0 V1+14 | | | IgM ELISA PfCSP OD 1.0 V3 | | | IgM ELISA PfCSP OD 1.0 V3+14 | | | IgM ELISA PfCSP OD 1.0 V3+28 | | | IgM ELISA PfCSP OD 1.0 V3+84 | | |
| | | | | | V1 | V1+14 | Net | Ratio | V3 | Net | Ratio | V3+14 | Net | Ratio | V3+28 | Net | Ratio | V3+84 | Net | Ratio |
| 1 | 20 - 50 | 3x $2.0x10^5$ | BFSPZL-008 | PfSPZ-LARC2 Vaccine | 365 | 388 | 23 | 1.06 | 711 | 346 | 1.95 | 724 | 359 | 1.98 | 2354 | 1989 | 6.45 | 566 | 201 | 1.55 |
| 1 | 20 - 50 | 3x $2.0x10^5$ | BFSPZL-013 | PfSPZ-LARC2 Vaccine | 464 | 432 | 1 | 1.00 | 396 | 1 | 1.00 | 130 | 1 | 1.00 | 257 | 1 | 1.00 | 171 | 1 | |
| 1 | 20 - 50 | 3x $2.0x10^5$ | BFSPZL-015 | PfSPZ-LARC2 Vaccine | 654 | 455 | 1 | 1.00 | 832 | 178 | 1.27 | 892 | 238 | 1.36 | 851 | 197 | 1.30 | 948 | 294 | 1.45 |
| 1 | 20 - 50 | 3x $2.0x10^5$ | BFSPZL-017 | PfSPZ-LARC2 Vaccine | 568 | 507 | 1 | 1.00 | 356 | 1 | 1.00 | 528 | 1 | 1.00 | 373 | 1 | 1.00 | 258 | 1 | 1.00 |
| 1 | 20 - 50 | 3x $2.0x10^5$ | BFSPZL-025 | PfSPZ-LARC2 Vaccine | 654 | 475 | 1 | 1.00 | 832 | 178 | 1.27 | 908 | 254 | 1.39 | 851 | 197 | 1.30 | NS | NS | NS |
| 1 | 20 - 50 | 3x $2.0x10^5$ | BFSPZL-036 | PfSPZ-LARC2 Vaccine | 744 | 895 | 151 | 1.20 | 1014 | 270 | 1.36 | 839 | 95 | 1.13 | 1821 | 1077 | 2.45 | 695 | 1 | 1.00 |
| 1 | 20 - 50 | 3x $2.0x10^5$ | BFSPZL-037 | PfSPZ-LARC2 Vaccine | 228 | 568 | 340 | 2.49 | 507 | 279 | 2.22 | 437 | 209 | 1.92 | 318 | 90 | 1.39 | 374 | 146 | 1.64 |
| 1 | 20 - 50 | 3x $2.0x10^5$ | BFSPZL-039 | PfSPZ-LARC2 Vaccine | 449 | 296 | 1 | 1.00 | 379 | 1 | 1.00 | 703 | 254 | 1.57 | 423 | 1 | 1.00 | 788 | 339 | 1.76 |
| 1 | 20 - 50 | 3x $2.0x10^5$ | BFSPZL-056 | PfSPZ-LARC2 Vaccine | 948 | 207 | 1 | 1.00 | 204 | 1 | 1.00 | 522 | 1 | 1.00 | 212 | 1 | 1.00 | 259 | 1 | 1.00 |
| 1 | 20 - 50 | 3x $2.0x10^5$ | BFSPZL-067 | PfSPZ-LARC2 Vaccine | 258 | 476 | 218 | 1.84 | 566 | 308 | 2.19 | 374 | 116 | 1.45 | 474 | 216 | 1.84 | 688 | 430 | 2.67 |
| **Median** | | | | | **516** | **465** | **1** | **1** | **537** | **178** | **1.27** | **616** | **163** | **1.38** | **449** | **144** | **1.30** | **566** | **146** | **1.50** |
| **Mean** | | | | | **533** | **470** | **74** | **1.26** | **580** | **156** | **1.43** | **606** | **153** | **1.38** | **793** | **377** | **1.87** | **527** | **157** | **1.51** |
| **GeoMean** | | | | | **488** | **440** | **7** | **1.19** | **524** | **28** | **1.36** | **538** | **41** | **1.34** | **576** | **33** | **1.53** | **457** | **22** | **1.43** |
| 1 | 20 - 50 | 3x NS | BFSPZL-014 | Normal Saline placebo | 630 | 888 | 258 | 1.41 | 536 | 1 | 1.00 | 638 | 8 | 1.01 | 836 | 206 | 1.33 | 1398 | 768 | 2.22 |
| 1 | 20 - 50 | 3x NS | BFSPZL-016 | Normal Saline placebo | 392 | 152 | 1 | 1.00 | 302 | 1 | 1.00 | 170 | 1 | 1.00 | 189 | 1 | 1.00 | 836 | 444 | 2.13 |
| 1 | 20- 50 | 3x NS | BFSPZL-023 | Normal Saline placebo | 207 | 204 | 1 | 1.00 | 178 | 1 | 1.00 | 212 | 5 | 1.02 | 259 | 52 | 1.25 | 228 | 21 | 1.10 |
| 1 | 20 - 50 | 3x NS | BFSPZL-027 | Normal Saline placebo | 476 | 566 | 90 | 1.19 | 439 | 1 | 1.00 | 474 | 1 | 1.00 | 688 | 212 | 1.45 | 449 | 1 | 1.00 |
| 1 | 20 - 50 | 3x NS | BFSPZL-032 | Normal Saline placebo | 296 | 379 | 83 | 1.28 | 774 | 478 | 2.61 | 423 | 127 | 1.43 | 788 | 492 | 2.66 | 661 | 365 | 2.23 |
| **Median** | | | | | **3,005** | **2,897** | **1** | **1** | **439** | **1** | **1.00** | **423** | **5** | **1.01** | **688** | **206** | **1.33** | **661** | **365** | **2.13** |
| **Mean** | | | | | **2,487** | **2,390** | **18** | **1** | **446** | **96** | **1.32** | **383** | **28** | **1.09** | **552** | **193** | **1.54** | **714** | **320** | **1.74** |
| **GeoMean** | | | | | **1,963** | **1,769** | **5** | **1** | **396** | **3** | **1.21** | **341** | **6** | **1.08** | **467** | **65** | **1.45** | **602** | **76** | **1.63** |

### B. <u>Groups 2-5 – PfCSP IgM</u>

| Group | Age | Dose | ID | Treatment Arm Allocation | IgM ELISA PfCSP OD 1.0 | IgM ELISA PfCSP OD 1.0 V1+14 | | | IgM ELISA PfCSP OD 1.0 V1+28 | | | IgM ELISA PfCSP OD 1.0 V1+84 | | |
|---|---|---|---|---|---|---|---|---|---|---|---|---|---|---|
| | | | | | **V1** | **V1+14** | **Net** | **Ratio** | **V1+28** | **Net** | **Ratio** | **V1+84** | **Net** | **Ratio** |
| 2 | 20 - 50 | 1x 4.0x$10^5$ | BFSPZL-011 | PfSPZ-LARC2 Vaccine | 783 | 558 | 1 | 1.00 | 658 | 1 | 1.00 | 519 | 1 | 1.00 |
| 2 | 20 - 50 | 1x 4.0x$10^5$ | BFSPZL-012 | PfSPZ-LARC2 Vaccine | 1067 | 991 | 1 | 1.00 | 1101 | 34 | 1.03 | 1005 | 1 | 1.00 |
| 2 | 20 - 50 | 1x 4.0x$10^5$ | BFSPZL-029 | PfSPZ-LARC2 Vaccine | 267 | 649 | 382 | 2.43 | 695 | 428 | 2.60 | 678 | 411 | 2.54 |
| 2 | 20 - 50 | 1x 4.0x$10^5$ | BFSPZL-049 | PfSPZ-LARC2 Vaccine | 2253 | 1506 | 1 | 1.00 | 1302 | 1 | 1.00 | 1462 | 1 | 1.00 |
| 2 | 20 - 50 | 1x 4.0x$10^5$ | BFSPZL-058 | PfSPZ-LARC2 Vaccine | 689 | 634 | 1 | 1.00 | 813 | 124 | 1.18 | 806 | 117 | 1.17 |
| 2 | 20 - 50 | 1x 4.0x$10^5$ | BFSPZL-062 | PfSPZ-LARC2 Vaccine | 565 | 848 | 283 | 1.50 | 926 | 361 | 1.64 | 882 | 317 | 1.56 |
| 2 | 20 - 50 | 1x 4.0x$10^5$ | BFSPZL-063 | PfSPZ-LARC2 Vaccine | 998 | 1212 | 214 | 1.21 | 1041 | 43 | 1.04 | 1256 | 258 | 1.26 |
| 2 | 20 - 50 | 1x 4.0x$10^5$ | BFSPZL-066 | PfSPZ-LARC2 Vaccine | 514 | 684 | 170 | 1.33 | 801 | 287 | 1.56 | NS | NS | NS |
| 2 | 20 - 50 | 1x 4.0x$10^5$ | BFSPZL-069 | PfSPZ-LARC2 Vaccine | 746 | 617 | 1 | 1.00 | 765 | 19 | 1.03 | 750 | 4 | 1.01 |
| 2 | 20 - 50 | 1x 4.0x$10^5$ | BFSPZL-070 | PfSPZ-LARC2 Vaccine | 1501 | 2392 | 891 | 1.59 | 3020 | 1519 | 2.01 | 1559 | 58 | 1.04 |
| **Median** | | | | | **765** | **766** | **86** | **1.11** | **870** | **84** | **1.11** | **882** | **58** | **1.04** |
| **Mean** | | | | | **938** | **1009** | **195** | **1.31** | **1112** | **282** | **1.41** | **991** | **130** | **1.29** |
| **GeoMean** | | | | | **804** | **902** | **18** | **1.25** | **995** | **55** | **1.33** | **934** | **21** | **1.22** |
| 2 | 20 - 50 | 1x NS | BFSPZL-024 | Normal Saline placebo | 2437 | 1993 | 1 | 1.00 | 2211 | 1 | 1.00 | 1087 | 1 | 1.00 |
| 2 | 20 - 50 | 1x NS | BFSPZL-034 | Normal Saline placebo | 308 | 299 | 1 | 1.00 | 555 | 247 | 1.80 | 334 | 26 | 1.08 |
| 2 | 20 - 50 | 1x NS | BFSPZL-052 | Normal Saline placebo | 1061 | 1291 | 230 | 1.22 | 1680 | 619 | 1.58 | 1513 | 452 | 1.43 |
| 2 | 20 - 50 | 1x NS | BFSPZL-061 | Normal Saline placebo | 1055 | 2322 | 1267 | 2.20 | 1248 | 193 | 1.18 | 933 | 1 | 1.00 |
| 2 | 20 - 50 | 1x NS | BFSPZL-068 | Normal Saline placebo | 1337 | 1887 | 550 | 1.41 | 1158 | 1 | 1.00 | 1394 | 57 | 1.04 |
| **Median** | | | | | **1061** | **1887** | **230** | **1.22** | **1248** | **193** | **1.18** | **1087** | **26** | **1.04** |
| **Mean** | | | | | **1240** | **1558** | **410** | **1.37** | **1370** | **212** | **1.31** | **1052** | **107** | **1.11** |
| **GeoMean** | | | | | **1024** | **1275** | **44** | **1.30** | **1244** | **31** | **1.28** | **935** | **15** | **1.10** |
| 3 | 11 – 19 | 1x 2.0x$10^5$ | BFSPZL-095 | PfSPZ-LARC2 Vaccine | 1442 | 1063 | 1 | 1.00 | NS | NS | NS | NS | NS | NS |
| 3 | 11 – 19 | 1x 2.0x$10^5$ | BFSPZL-099 | PfSPZ-LARC2 Vaccine | 851 | 2841 | 1990 | 3.34 | 925 | 74 | 1.09 | 696 | 1 | 1.00 |
| 3 | 11 – 19 | 1x 2.0x$10^5$ | BFSPZL-101 | PfSPZ-LARC2 Vaccine | 431 | 937 | 506 | 2.17 | 420 | 1 | 1.00 | NS | NS | NS |
| 3 | 11 – 19 | 1x 2.0x$10^5$ | BFSPZL-105 | PfSPZ-LARC2 Vaccine | 862 | 699 | 1 | 1.00 | 697 | 1 | 1.00 | 766 | 1 | 1.00 |
| 3 | 11 – 19 | 1x 2.0x$10^5$ | BFSPZL-108 | PfSPZ-LARC2 Vaccine | 940 | 8853 | 7913 | 9.42 | 1075 | 135 | 1.14 | 503 | 1 | 1.00 |
| 3 | 11 – 19 | 1x 2.0x$10^5$ | BFSPZL-110 | PfSPZ-LARC2 Vaccine | 1120 | 1899 | 779 | 1.70 | 1403 | 283 | 1.25 | 1017 | 1 | 1.00 |
| 3 | 11 – 19 | 1x 2.0x$10^5$ | BFSPZL-114 | PfSPZ-LARC2 Vaccine | 615 | 506 | 1 | 1.00 | 383 | 1 | 1.00 | 500 | 1 | 1.00 |
| 3 | 11 – 19 | 1x 2.0x$10^5$ | BFSPZL-115 | PfSPZ-LARC2 Vaccine | 1717 | 1212 | 1 | 1.00 | 656 | 1 | 1.00 | 682 | 1 | 1.00 |
| 3 | 11 – 19 | 1x 2.0x$10^5$ | BFSPZL-118 | PfSPZ-LARC2 Vaccine | 1029 | 2356 | 1327 | 2.29 | 983 | 1 | 1.00 | 848 | 1 | 1.00 |
| 3 | 11 – 19 | 1x 2.0x$10^5$ | BFSPZL-121 | PfSPZ-LARC2 Vaccine | 1100 | 1137 | 37 | 1.03 | 1093 | 1 | 1.00 | 1035 | 1 | 1.00 |
| **Median** | | | | | **985** | **1175** | **272** | **1.36** | **925** | **1** | **1.00** | **731** | **1** | **1.00** |
| **Mean** | | | | | **1011** | **2150** | **1256** | **2.39** | **848** | **55** | **1.05** | **756** | **1** | **1.00** |
| **GeoMean** | | | | | **946** | **1492** | **56** | **1.75** | **783** | **5** | **1.05** | **731** | **1** | **1.00** |
| 3 | 11 – 19 | 1x NS | BFSPZL-089 | Normal Saline placebo | 2984 | 1502 | 1 | 1.00 | 4489 | 1505 | 1.50 | 1373 | 1 | 1.00 |
| 3 | 11 – 19 | 1x NS | BFSPZL-103 | Normal Saline placebo | 1475 | 2558 | 1083 | 1.73 | 1518 | 43 | 1.03 | 1165 | 1 | 1.00 |
| 3 | 11 – 19 | 1x NS | BFSPZL-107 | Normal Saline placebo | 1811 | 3405 | 1594 | 1.88 | 1644 | 1 | 1.00 | 1216 | 1 | 1.00 |
| 3 | 11 – 19 | 1x NS | BFSPZL-112 | Normal Saline placebo | 1922 | 1636 | 1 | 1.00 | 1699 | 1 | 1.00 | 1744 | 1 | 1.00 |
| 3 | 11 – 19 | 1x NS | BFSPZL-116 | Normal Saline placebo | 274 | 209 | 1 | 1.00 | 296 | 22 | 1.08 | 331 | 57 | 1.21 |
| **Median** | | | | | **1811** | **1636** | **1** | **1.00** | **1644** | **22** | **1.03** | **1216** | **1** | **1.00** |
| **Mean** | | | | | **1693** | **1862** | **536** | **1.32** | **1929** | **314** | **1.12** | **1166** | **12** | **1.04** |
| **GeoMean** | | | | | **1332** | **1349** | **18** | **1.27** | **1413** | **17** | **1.11** | **1023** | **2** | **1.04** |

Note: the spanning header "IgM ELISA PfCSP OD 1.0" is printed over all result columns.

| | | | | | | | | | | | | | | |
|---|---|---|---|---|---|---|---|---|---|---|---|---|---|---|
| 4 | 6 - 10 | 1x 2.0x$10^5$ | BFSPZL-073 | PfSPZ-LARC2 Vaccine | 2766 | 3083 | 317 | 1.11 | 1759 | 1 | 1.00 | 3537 | 771 | 1.28 |
| 4 | 6 - 10 | 1x 2.0x$10^5$ | BFSPZL-075 | PfSPZ-LARC2 Vaccine | 1626 | 2638 | 1012 | 1.62 | 1946 | 320 | 1.20 | 2391 | 765 | 1.47 |
| 4 | 6 - 10 | 1x 2.0x$10^5$ | BFSPZL-081 | PfSPZ-LARC2 Vaccine | 937 | 2458 | 1521 | 2.62 | 1406 | 469 | 1.50 | NS | NS | NS |
| 4 | 6 - 10 | 1x 2.0x$10^5$ | BFSPZL-082 | PfSPZ-LARC2 Vaccine | 511 | 626 | 115 | 1.23 | 618 | 107 | 1.21 | 1174 | 663 | 2.30 |
| 4 | 6 - 10 | 1x 2.0x$10^5$ | BFSPZL-084 | PfSPZ-LARC2 Vaccine | 477 | 1012 | 535 | 2.12 | 545 | 68 | 1.14 | 1205 | 728 | 2.53 |
| 4 | 6 - 10 | 1x 2.0x$10^5$ | BFSPZL-085 | PfSPZ-LARC2 Vaccine | 991 | 1844 | 853 | 1.86 | 991 | 1 | 1.00 | 1324 | 333 | 1.34 |
| 4 | 6 - 10 | 1x 2.0x$10^5$ | BFSPZL-088 | PfSPZ-LARC2 Vaccine | 548 | 1714 | 1166 | 3.13 | 1048 | 500 | 1.91 | 1222 | 674 | 2.23 |
| 4 | 6 - 10 | 1x 2.0x$10^5$ | BFSPZL-090 | PfSPZ-LARC2 Vaccine | 694 | 888 | 194 | 1.28 | 605 | 1 | 1.00 | 904 | 210 | 1.30 |
| 4 | 6 - 10 | 1x 2.0x$10^5$ | BFSPZL-119 | PfSPZ-LARC2 Vaccine | 2010 | 3985 | 1975 | 1.98 | 2639 | 629 | 1.31 | 1748 | 1 | 1.00 |
| 4 | 6 - 10 | 1x 2.0x$10^5$ | BFSPZL-120 | PfSPZ-LARC2 Vaccine | 485 | 396 | 1 | 1.00 | 387 | 1 | 0.80 | 630 | 145 | 1.30 |
| **Median** | | | | | **816** | **1779** | **694** | **1.74** | **1020** | **88** | **1.17** | **1222** | **663** | **1.34** |
| **Mean** | | | | | **1105** | **1864** | **769** | **1.80** | **1194** | **210** | **1.21** | **1571** | **477** | **1.64** |
| **GeoMean** | | | | | **907** | **1496** | **321** | **1.68** | **1004** | **28** | **1.17** | **1389** | **232** | **1.56** |
| 4 | 6 - 10 | 1x NS | BFSPZL-074 | Normal Saline placebo | 1744 | 1754 | 10 | 1.01 | 1689 | 1 | 1.00 | 2135 | 391 | 1.22 |
| 4 | 6 - 10 | 1x NS | BFSPZL-079 | Normal Saline placebo | 1032 | 766 | 1 | 1.00 | 516 | 1 | 1.00 | 802 | 1 | 1.00 |
| 4 | 6 - 10 | 1x NS | BFSPZL-080 | Normal Saline placebo | 1407 | 1484 | 77 | 1.05 | 1191 | 1 | 1.00 | 2773 | 1366 | 1.97 |
| 4 | 6 - 10 | 1x NS | BFSPZL-083 | Normal Saline placebo | 1387 | 1976 | 589 | 1.42 | 1063 | 1 | 1.00 | 2390 | 1003 | 1.72 |
| 4 | 6 - 10 | 1x NS | BFSPZL-087 | Normal Saline placebo | 610 | 825 | 215 | 1.35 | 620 | 10 | 1.02 | 867 | 257 | 1.42 |
| **Median** | | | | | **1387** | **1484** | **77** | **1.05** | **1063** | **1** | **1.00** | **2135** | **391** | **1.42** |
| **Mean** | | | | | **1236** | **1361** | **178** | **1.17** | **1016** | **3** | **1.00** | **1793** | **604** | **1.47** |
| **GeoMean** | | | | | **1165** | **1266** | **40** | **1.15** | **927** | **2** | **1.00** | **1580** | **169** | **1.43** |
| 5 | 1 - 5 | 1x 2.0x$10^5$ | BFSPZL-122 | PfSPZ-LARC2 Vaccine | 572 | 1160 | 588 | 2.03 | 1280 | 708 | 2.24 | 231 | 1 | 1.00 |
| 5 | 1 - 5 | 1x 2.0x$10^5$ | BFSPZL-130 | PfSPZ-LARC2 Vaccine | 857 | 1022 | 165 | 1.19 | 855 | 1 | 1.00 | 218 | 1 | 1.00 |
| 5 | 1 - 5 | 1x 2.0x$10^5$ | BFSPZL-131 | PfSPZ-LARC2 Vaccine | 1237 | 685 | 1 | 1.00 | 564 | 1 | 1.00 | 1 | 1 | 1.00 |
| 5 | 1 - 5 | 1x 2.0x$10^5$ | BFSPZL-132 | PfSPZ-LARC2 Vaccine | 1677 | 2265 | 588 | 1.35 | 1008 | 1 | 1.00 | 248 | 1 | 1.00 |
| 5 | 1 - 5 | 1x 2.0x$10^5$ | BFSPZL-134 | PfSPZ-LARC2 Vaccine | 1957 | 2513 | 556 | 1.28 | 1161 | 1 | 1.00 | 67 | 1 | 1.00 |
| 5 | 1 - 5 | 1x 2.0x$10^5$ | BFSPZL-136 | PfSPZ-LARC2 Vaccine | 2586 | 3298 | 712 | 1.28 | 1797 | 1 | 1.00 | 76 | 1 | 1.00 |
| 5 | 1 - 5 | 1x 2.0x$10^5$ | BFSPZL-141 | PfSPZ-LARC2 Vaccine | 1017 | 1514 | 497 | 1.49 | 722 | 1 | 1.00 | 33 | 1 | 1.00 |
| 5 | 1 - 5 | 1x 2.0x$10^5$ | BFSPZL-144 | PfSPZ-LARC2 Vaccine | 1119 | 975 | 1 | 1.00 | 585 | 1 | 1.00 | 88 | 1 | 1.00 |
| 5 | 1 - 5 | 1x 2.0x$10^5$ | BFSPZL-146 | PfSPZ-LARC2 Vaccine | 622 | 631 | 9 | 1.01 | 638 | 16 | 1.03 | 34 | 1 | 1.00 |
| 5 | 1 - 5 | 1x 2.0x$10^5$ | BFSPZL-147 | PfSPZ-LARC2 Vaccine | 1313 | 4433 | 3120 | 3.38 | 1481 | 168 | 1.13 | 81 | 1 | 1.00 |
| **Median** | | | | | **1178** | **1337** | **527** | **1.28** | **932** | **1** | **1.00** | **79** | **1** | **1.00** |
| **Mean** | | | | | **1296** | **1850** | **624** | **1.50** | **1009** | **90** | **1.14** | **108** | **1** | **1.00** |
| **GeoMean** | | | | | **1169** | **1513** | **112** | **1.39** | **936** | **4** | **1.10** | **59** | **1** | **1.00** |
| 5 | 1 - 5 | 1x NS | BFSPZL-124 | Normal Saline placebo | 1068 | 1188 | 120 | 1.11 | 833 | 1 | 1.00 | 66 | 1 | 1.00 |
| 5 | 1 - 5 | 1x NS | BFSPZL-135 | Normal Saline placebo | 1011 | 684 | 1 | 1.00 | 501 | 1 | 1.00 | 81 | 1 | 1.00 |
| 5 | 1 - 5 | 1x NS | BFSPZL-140 | Normal Saline placebo | 1176 | 977 | 1 | 1.00 | 701 | 1 | 1.00 | 88 | 1 | 1.00 |
| 5 | 1 - 5 | 1x NS | BFSPZL-143 | Normal Saline placebo | 831 | 516 | 1 | 1.00 | 500 | 1 | 1.00 | 96 | 1 | 1.00 |
| 5 | 1 - 5 | 1x NS | BFSPZL-149 | Normal Saline placebo | 1422 | 3198 | 1776 | 2.25 | 2013 | 591 | 1.42 | 71 | 1 | 1.00 |
| **Median** | | | | | **1068** | **977** | **1** | **1.00** | **701** | **1** | **1.00** | **81** | **1** | **1.00** |
| **Mean** | | | | | **1102** | **1313** | **380** | **1.27** | **910** | **119** | **1.08** | **80** | **1** | **1.00** |
| **GeoMean** | | | | | **1085** | **1056** | **12** | **1.20** | **783** | **4** | **1.07** | **80** | **1** | **1.00** |

### C. Groups 1 – PfCSP IgA

| Group | Age | Dose | ID | Treatment Arm Allocation | IgA ELISA PfCSP OD 1.0 | | | | | | | | | | | | | | | |
|---|---|---|---|---|---|---|---|---|---|---|---|---|---|---|---|---|---|---|---|---|
| | | | | | IgA ELISA PfCSP OD 1.0 | IgA ELISA PfCSP OD 1.0 V1+14 | | | IgA ELISA PfCSP OD 1.0 V3 | | | IgA ELISA PfCSP OD 1.0 V3+14 | | | IgA ELISA PfCSP OD 1.0 V3+28 | | | IgA ELISA PfCSP OD 1.0 V3+84 | | |
| | | | | | **V1** | **V1+14** | **Net** | **Ratio** | **V3** | **Net** | **Ratio** | **V3+14** | **Net** | **Ratio** | **V3+28** | **Net** | **Ratio** | **V3+84** | **Net** | **Ratio** |
| 1 | 20 - 50 | 3x 2.0x10$^5$ | BFSPZL-008 | PfSPZ-LARC2 Vaccine | 258 | 1 | 1 | 1.00 | 1 | 1 | 1.00 | 1 | 1 | 1.00 | 1 | 1 | 1.00 | 247 | 1 | 1.00 |
| 1 | 20 - 50 | 3x 2.0x10$^5$ | BFSPZL-013 | PfSPZ-LARC2 Vaccine | 1 | 1 | 1 | 1.00 | 272 | 271 | 272.00 | 275 | 274 | 275.00 | 73 | 72 | 73.00 | 1 | 1 | 1.00 |
| 1 | 20 - 50 | 3x 2.0x10$^5$ | BFSPZL-015 | PfSPZ-LARC2 Vaccine | 99 | 130 | 31 | 1.31 | 596 | 497 | 6.02 | 323 | 224 | 3.26 | 2211 | 2112 | 22.33 | 582 | 483 | 5.88 |
| 1 | 20 - 50 | 3x 2.0x10$^5$ | BFSPZL-017 | PfSPZ-LARC2 Vaccine | 509 | 271 | 1 | 1.00 | 158 | 1 | 1.00 | 228 | 1 | 1.00 | 709 | 200 | 1.39 | 238 | 1 | 1.00 |
| 1 | 20 - 50 | 3x 2.0x10$^5$ | BFSPZL-025 | PfSPZ-LARC2 Vaccine | 406 | 429 | 23 | 1.06 | 827 | 421 | 2.04 | 1065 | 659 | 2.62 | 399 | 1 | 1.00 | 509 | 103 | 1.25 |
| 1 | 20 - 50 | 3x 2.0x10$^5$ | BFSPZL-036 | PfSPZ-LARC2 Vaccine | 156 | 277 | 121 | 1.78 | 132 | 1 | 1.00 | 203 | 47 | 1.30 | 116 | 1 | 1.00 | 202 | 46 | 1.29 |
| 1 | 20 - 50 | 3x 2.0x10$^5$ | BFSPZL-037 | PfSPZ-LARC2 Vaccine | 824 | 163 | 1 | 1.00 | 179 | 1 | 1.00 | 275 | 1 | 1.00 | 233 | 1 | 1.00 | 291 | 1 | 1.00 |
| 1 | 20 - 50 | 3x 2.0x10$^5$ | BFSPZL-039 | PfSPZ-LARC2 Vaccine | 394 | 344 | 1 | 1.00 | 970 | 576 | 2.46 | 463 | 69 | 1.18 | 327 | 1 | 1.00 | 307 | 1 | 1.00 |
| 1 | 20 - 50 | 3x 2.0x10$^5$ | BFSPZL-056 | PfSPZ-LARC2 Vaccine | 397 | 456 | 59 | 1.15 | 383 | 1 | 1.00 | 508 | 111 | 1.28 | 570 | 173 | 1.44 | 281 | 1 | 1.00 |
| 1 | 20 - 50 | 3x 2.0x10$^5$ | BFSPZL-067 | PfSPZ-LARC2 Vaccine | 828 | 585 | 1 | 1.00 | 590 | 1 | 1.00 | 600 | 1 | 1.00 | 592 | 1 | 1.00 | 734 | 1 | 1.00 |
| **Median** | | | | | **396** | **274** | **1** | **1.00** | **328** | **1** | **1.00** | **299** | **58** | **1.23** | **363** | **1** | **1.00** | **286** | **1** | **1.00** |
| **Mean** | | | | | **387** | **266** | **24** | **1.13** | **411** | **177** | **28.85** | **394** | **139** | **28.86** | **523** | **256** | **10.42** | **339** | **64** | **1.54** |
| **GeoMean** | | | | | **197** | **95** | **5** | **1.11** | **201** | **11** | **2.46** | **210** | **21** | **2.32** | **206** | **9** | **2.25** | **191** | **4** | **1.25** |
| 1 | 20 - 50 | 3x NS | BFSPZL-014 | Normal Saline placebo | 13 | 1 | 1 | 1.00 | 2 | 1 | 1.00 | 1 | 1 | 1.00 | 1 | 1 | 1.00 | 1 | 1 | 1.00 |
| 1 | 20 - 50 | 3x NS | BFSPZL-016 | Normal Saline placebo | 1 | 1 | 1 | 1.00 | 1 | 1 | 1.00 | 86 | 85 | 86.00 | 55 | 54 | 55.00 | 21 | 20 | 21.00 |
| 1 | 20- 50 | 3x NS | BFSPZL-023 | Normal Saline placebo | 334 | 440 | 106 | 1.32 | 271 | 1 | 1.00 | 479 | 145 | 1.43 | 419 | 85 | 1.25 | 400 | 66 | 1.20 |
| 1 | 20 - 50 | 3x NS | BFSPZL-027 | Normal Saline placebo | 287 | 810 | 523 | 2.82 | 206 | 1 | 1.00 | 219 | 1 | 1.00 | 430 | 143 | 1.50 | 827 | 540 | 2.88 |
| 1 | 20 - 50 | 3x NS | BFSPZL-032 | Normal Saline placebo | 201 | 212 | 11 | 1.05 | 850 | 649 | 4.23 | 307 | 106 | 1.53 | 371 | 170 | 1.85 | 605 | 404 | 3.01 |
| **Median** | | | | | **201** | **212** | **11** | **1.05** | **206** | **1** | **1.00** | **219** | **85** | **1.43** | **371** | **85** | **1.50** | **400** | **66** | **2.88** |
| **Mean** | | | | | **167** | **293** | **128** | **1.44** | **266** | **131** | **1.65** | **218** | **68** | **18.19** | **255** | **91** | **12.12** | **371** | **206** | **5.82** |
| **GeoMean** | | | | | **83** | **110** | **23** | **1.33** | **100** | **4** | **1.33** | **227** | **41** | **3.06** | **267** | **99** | **3.10** | **279** | **114** | **3.63** |
